# Immune profiling identifies predictive biomarkers and highlights the potential efficacy of IL-6R blockade in checkpoint inhibitor–related myocarditis

**DOI:** 10.1101/2025.01.16.25320675

**Authors:** Sarah Boughdad, Douglas Daoudlarian, Robin Bartolini, Sofiya Latifyan, Jacqueline Doms, Hasna Bouchaab, Karim Abdelhamid, Nabila Ferahta, Nuria Mederos, Victor Joo, Antonia Stamatiou, Lucrezia Mencarelli, Nicolas Etienne, Khalil Zaman, Matthieu Perreau, Craig Fenwick, Keyvan Shabafrouz, Giuseppe Pantaleo, Solange Peters, Michel Obeid

## Abstract

**Background and aims:** Immune checkpoint inhibitor–associated myocarditis (ICI-My) is a rare but potentially life-threatening complication. Advancing our understanding of its underlying immunological mechanisms is essential for the development of improved diagnostic tools and targeted treatment strategies, with the aim of ultimately enhancing patient outcomes and minimizing morbidity and mortality.

**Methods:** This retrospective single-center study (July 2019–June 2024) identified 33 patients who developed ICI-My. A Comprehensive immuno-profiling was conducted using 49 cytokines, 7 traditional cardiac biomarkers, and 46 mass cytometry markers. These profiles were compared to the baseline levels of a cohort of 97 cancer patients prior to ICI treatment. The analysis assessed the identification of biomarkers for differentiating low and high-grade myocarditis and corticosteroids (CS)-refractory ICI-My. The therapeutic efficacy of tocilizumab (anti-IL6R) was additionally assessed in seven cases of CS-refractory myocarditis.

**Results:** ICI-My patients showed marked elevations in IL-6, CXCL9, CXCL10, CXCL13, VEGF-A, and sCD25 compared with baseline cancer patients prior to ICI initiation. High-grade myocarditis was characterized by lower levels of CCL4 and CXCL12, with predictive accuracies of 78.6% and 82.1%, respectively. In contrast, conventional biomarkers (cTnT, cTnI, CK, CK-MB, NT-ProBNP, and d-dimers) failed to differentiate disease severity. Mass cytometry revealed a distinct immune profile in ICI-My, including increased immature neutrophils, reduced switched and unswitched memory B cells, elevated double-positive (CD38⁺/HLA-DR⁺) T cells across CD4⁺ and CD8⁺ subsets, decreased CXCR5⁺ leukocytes, and diminished CXCR3 expression within all memory T-cell subsets. Notably, no complement activation was detected. HGF, CXCL10, and BDNF successfully discriminated patients requiring immunosuppression from those untreated (accuracies of 89%, 79%, and 79%, respectively), while IL-18 and CCL4 predicted the need for tocilizumab (TCZ) therapy (accuracies of 79% and 82%, respectively). This underscores the dual benefit of CCL4. In addition, all cases of corticosteroid (CS)-refractory myocarditis (n=8), including those unresponsive to mycophenolate mofetil (MMF) or infliximab, responded effectively to TCZ.

**Conclusions:** This study provides the first comprehensive immuno-profile of ICI-My, revealing distinct cytokine signatures and immune cell alterations associated with disease severity. CCL4 and CXCL12 outperformed traditional cardiac biomarkers as prognostic tools, while IL-18 and CCL4 emerged as key predictors for tocilizumab therapy, which could offer a personalized therapeutic approach. The absence of complement activation indicates that cytokine-mediated and cellular pathways are central to ICI-My pathogenesis. Notably, the success of anti-IL-6 therapy in corticosteroid-refractory cases highlights new therapeutic opportunities, enhancing patient care and guiding future interventions.

**Key question:** How can immune profiling and the identification of predictive biomarkers enhance the diagnosis, risk stratification, and treatment of immune checkpoint inhibitor–related myocarditis (ICI-My)?

**Key findings:**

1. **Cytokine-mediated pathogenesis**. Patients with ICI-My exhibit significantly elevated levels of several pro-inflammatory cytokines, including IL-6, CXCL9, CXCL10, CXCL13, VEGF-A, and sCD25, highlighting a strong cytokine-driven inflammatory response.
2. **Immune cell alterations**. Shifts in T-cell populations and neutrophil subsets highlight a central role for T-cell–mediated mechanisms in the pathogenesis of ICI-My.
3. **No complement activation**. The absence of complement activation further supports the dominance of cytokine- and T-cell–mediated pathways over complement-driven processes in ICI-My.
4. **Superior biomarkers for risk stratification**. CCL4 and CXCL12 outperform traditional cardiac markers (e.g., troponins, NT-proBNP) in differentiating high-versus low-grade myocarditis, offering improved risk stratification.
5. **Biomarkers for immunosuppression decisions**. IL-18, CCL2 and CXCL13 accurately distinguish patients needing immunosuppression from those who do not, with PPV of 100%.
6. **Predictive biomarkers for tocilizumab**. IL-18 and CCL4 predict the need for TCZ (anti–IL-6 receptor) in refractory cases. Notably, CCL4 confers a dual benefit: it not only contributes stratifying disease severity but also helps discriminating patients most likely to benefit from tocilizumab.
7. **Therapeutic efficacy of tocilizumab.** TCZ is effective in treating refractory ICI-My, including in patients unresponsive to other immunosuppressive agents such as anti-TNFα or MMF.

**Take home message:** This study underscores the pivotal importance of cytokine and immune cell profiling in the diagnosis and management of ICI-related myocarditis. Incorporating a concise biomarker panel— encompassing CCL4, CXCL12, IL-18, HGF, CXCL10, and BDNF—into routine clinical workflows could facilitate early risk stratification and guide therapeutic decisions. The results support the notion that ICI-My is predominantly driven by cytokine- and T-cell–mediated pathways, without involvement of the complement system. Furthermore, TCZ appears highly efficacious in patients who are unresponsive to corticosteroids or other immunosuppressive agents, solidifying its potential as a valuable therapeutic option. These findings pave the way for more targeted interventions aimed at mitigating severe cardiac toxicities while preserving the anti-tumor benefits of ICIs.

## Introduction

Among the various cardiac immune-related adverse events (irAEs), myocarditis is the most commonly reported and is associated with significant morbidity and mortality.^1,2^ Although considered rare (approximately 1% of monotherapy cases and 2–3% with combination regimens^1,2^), the true incidence may be underestimated ^3^. In addition, myocarditis with combined ICI tends to be more severe and carries an increased risk of death^4^. ICI-related myocarditis presents a significant clinical challenge due to its heterogeneous presentation, which frequently manifests as nonspecific symptoms such as fatigue, chest pain, and dyspnea, and its potential for rapid progression to severe cardiac dysfunction^5^. The current diagnostic modalities for ICI-related myocarditis (ICI-My) primarily rely on clinical evaluation, the measurement of biomarkers such as troponins and natriuretic peptides, and the use of imaging techniques including echocardiography and cardiac magnetic resonance imaging (CMRI)^6^. However, these approaches have limited sensitivity and specificity for accurate risk stratification and personalized management of affected patients^7^. The early diagnosis and treatment of ICI-My are of paramount importance in reducing the mortality rate. A recent study demonstrated that up to 50% of patients achieved better outcomes when diagnosed and treated promptly^8–10^.

In addition, the identification of circulating biomarkers that facilitate early diagnosis, assess disease severity, and predict treatment response is imperative for improving ICI-My management. ICI-My is characterized by increased infiltration of CD4⁺ and CD8⁺ T lymphocytes^11^, including T cells that target cardiac myosin^12,13^ and activated Temra (T-effector memory cells re-expressing CD45RA) CD8⁺ T-cells^14^. Recent studies in both murine models and patients with ICI-My have reported a significant enrichment of macrophages, particularly an inflammatory CCR2⁺ subpopulation that highly expresses CXCL9 and CXCL10, originating from CCR2⁺ monocytes^15^. Although serum cytokines were not assessed in this study, our team has previously reported in a small cohort elevated levels of pro-inflammatory cytokines, including CXCL9, CXCL10, IL-6 and CCL2^16^, which can induce monocyte chemotaxis to the myocardium via CCR2 receptor mediation. Despite these important advancements, considerable gaps remain in our understanding of the immunopathogenesis of ICI-My. Most existing studies have focused on a limited range of cytokines and conventional markers, potentially leading to an incomplete comprehension of the complex immune responses involved. A deeper exploration of the interplay between immune activation, the identification of predictive biomarkers, and the development of personalized therapeutic strategies is essential.

Advancements in immunological profiling, particularly the comprehensive analysis of cytokine networks and the application of mass cytometry, offer promising avenues for elucidating the intricate immune mechanisms underlying ICI-My and providing in-depth insight into the immune landscape^17^. By integrating these advanced immunoprofiling techniques with traditional biomarkers, it is possible to identify distinct immunological signatures associated with disease severity, treatment responsiveness, and long-term oncological outcomes.

In this retrospective study, we performed an in-depth immunoprofiling of patients with ICI-related myocarditis using a comprehensive set of 105 biomarkers, including 49 cytokines, chemokines and growth factors, 12 biological markers (classical cardiac biomarkers and complement analysis) and 44 cellular markers. The objectives of the study are as follows: to characterize the immunological signatures associated with the onset and severity of myocarditis, to identify a minimal set of predictive markers linked to disease severity and response to immunosuppressive treatments, and to explore the potential use of tocilizumab (TCZ) in treating refractory CS-myocarditis. The fulfillment of these objectives may improve the management of ICI-related myocarditis, thereby enhancing treatment safety by the development of personalized diagnostic and therapeutic interventions.

## Material and Methods

### Patient consent, ethical approval, and sample collection

This retrospective study (“Immuno-TOX“) was approved by the cantonal ethics committee, the Commission Cantonale d’Éthique de la Recherche sur l’Être Humain (CER-VD). All biological samples were collected during routine clinical practice, with no modifications to standard care protocols. All study participants provided informed consent for the research use of their data through the “consentement général“ process, which ensures coded data handling to maintain confidentiality. For patients who did not reply the “consentement général“ and also did not actively object to recruitment, enrollment was performed in compliance with Article 34 of the Swiss Federal Law on Human Research as authorized in the “Immuno-TOX” study.

### Pilot investigation, study design and patient cohort

This retrospective study conducted at the Services of Medical Oncology, Immunology, and Allergy Service of CHUV, included all cases of ICI-related myocarditis (ICI-My) diagnosed between January 2018 and June 2024. The cohort comprised 33 patients with cancer who had received ICIs and subsequently developed ICI-My. These patients continued to receive clinical follow-up care at the Lausanne Center for Immuno-Oncology Toxicities (LCIT Center), the Medical Oncology service, and, when necessary, with cardiology specialists. The principal objective of this study was to generate hypotheses rather than to draw definitive conclusions. The rarity of ICI-related myocarditis presents a significant challenge in assembling large cohorts, underscoring the need for external validation of our findings in independent and larger populations to enhance statistical power and generalizability. Notwithstanding these limitations, our study offers valuable real-world evidence that can inform future prospective research aimed at establishing robust evidence for the use of tocilizumab (TCZ) in managing ICI-related myocarditis (ICI-Myo). The presentation of these preliminary results is intended to inform clinical practice and encourage the scientific community to pursue further investigations in this area, with the ultimate goal of improving patient care through evidence-based interventions.

### Treatment of corticosteroid (CS)-refractory severe irCRS: scientific rationale and approach

The standard of care (SOC) for the management of CS-refractory severe ICI-My at our institution prioritizes the use of tocilizumab (TCZ). This approach minimizes the use of broad-spectrum immunosuppressive agents, such as etoposide, JAK inhibitors, abatacept and calcineurin inhibitors, which could potentially compromise the efficacy of ICIs. In alignment with the current ESMO guidelines^18^, which recommend TCZ for ICI-My, our SOC is further supported by previously published data from our small cohort, which demonstrates TCZ’s efficacy^16,19^. Despite the absence of large-scale cohort studies, we continue to employ TCZ to minimize immunosuppression, adhering to expert recommendations^20^. The protocol utilized in this retrospective study was developed based on preliminary data and clinical observations within our institution. We conducted a retrospective analysis of TCZ’s efficacy in treating seven (n=8) severe CS-refractory ICI-My across various tumor types, ICI regimens, and chemo-immunotherapy combinations. By validating its effectiveness, our objective is to reinforce TCZ as a cornerstone therapy for managing severe ICI-My and to strengthen its role in clinical practice.

### The diagnostic criteria for ICI-related myocarditis (ICI-My)

The diagnostic criteria for immune checkpoint inhibitor-related myocarditis (ICI-My) were based on the 2022 European Society of Cardiology (ESC) guidelines^6^, with severity assessed following American Society of Clinical Oncology (ASCO) guidelines^21^. All patients underwent comprehensive clinical evaluations and a thorough cardiologic work-up, which included measuring cardiac biomarkers such as serum cardiac troponin T (cTnT), serum cardiac troponin I (cTnI), creatine kinase (CK), CK-MB, NT-ProBNP, and D-dimers, along with performing a 12-lead electrocardiogram (ECG). According to the ESC 2022 guidelines^6^, the clinical diagnosis of ICI-related myocarditis was established by the presence of elevated cardiac troponins (cTnT or cTnI) in combination with either one major criterion or two minor criteria. In this study, the major criterion was a positive cardiac magnetic resonance (CMR) diagnosis of acute myocarditis based on the modified Lake Louise criteria^22^. The minor criteria included a clinical syndrome characterized by symptoms such as chest pain, shortness of breath, orthopnea, palpitations, syncope, cardiogenic shock, lower extremity edema, myalgias, muscle weakness, diplopia, ptosis, lightheadedness or dizziness, and fatigue; pathological ECG findings like new conduction system abnormalities or ventricular arrhythmias; decreased left ventricular function with or without regional wall motion abnormalities, excluding the Takotsubo pattern; associated ICI-related adverse events such as myositis, myopathy, or myasthenia gravis; and suggestive findings on CMR^6,22^. In a limited number of cases, endomyocardial biopsy (EMB) was performed to confirm the diagnosis of ICI-related myocarditis^23^. Additionally, in accordance with ESMO guidelines^18^, some patients underwent ^68^Ga-DOTATOC PET/CT metabolic imaging, particularly when CMR results were negative or unavailable. The decision to perform EMB was made after a careful assessment of each patient’s risk-benefit profile, ensuring that the diagnostic benefits outweighed the procedural risks, and was reserved for a limited number of ICI-My cases.

This comprehensive diagnostic approach enabled accurate identification and appropriate management of ICI-related myocarditis within the patient cohort.

### Immune profiling of circulating blood immune cells population by mass cytometry

Peripheral blood was stained following previously described protocols^17,24,25^. Briefly, cells were incubated with a 50 µL metal-conjugated antibody cocktail for 30 minutes at room temperature, subsequently washed, and fixed with 2.4% paraformaldehyde (PFA). A bulk lysis solution (Cytognos, Santa Marta de Tormes, Spain) was then applied for 15 minutes at room temperature to remove red blood cells, followed by incubation with additional metal-conjugated antibodies. The cells were labeled overnight at 4°C in 1.6% PFA containing 1 µM Cell-ID Intercalator (Standard BioTools, San Fransisco, USA). Data acquisition was performed using the HELIOS CyTOF system, with normalization conducted using EQ Four Element Calibration Beads via CyTOF software. Concurrently, pre-ICI samples from a large cohort of cancer patients (n = 97) were collected to assess pretreatment immune cell levels and distributions across various tumor types. The complete antibody panel and gating strategy are detailed. The complete 46 antibody panel and gating strategy are detailed in **Supplementary Figure 1**.

### Immune profiling of serum cytokine

As previously described^17,19,25–28^, we quantified serum concentrations of 49 cytokines, chemokines, and growth factors using the Luminex ProcartaPlex immunoassay panel. List of biomarkers, lower limit of detection and distribution of values within our cohort can be found in **Supplementary Table 1**. Samples values below or equal to the LLOD were replaced by the LLOD. Our study included a comparative analysis of two distinct cohorts, with cytokine levels assessed longitudinally at multiple time points: at the onset of ICI-related myocarditis and during subsequent immunosuppressive treatment. Additionally, pre-ICI serum samples from a large cohort of cancer patients (n=97) were collected to determine baseline cytokine levels across various tumor types. This approach enabled us to evaluate changes in cytokine profiles associated with ICI therapy and myocarditis development, providing insights into the immunological dynamics underlying these conditions.

### Statistical analysis

All statistical analysis in figures comparing cytokines were performed using GraphPad Prism 10.1.2 (GraphPad Software, Boston, Massachusetts USA). Mass cytometry data were analyzed in FlowJo. Statistical comparisons were made using Student’s t-test, Mann-Whitney U test, or Kruskal-Wallis test, as indicated. Significance was defined as p < 0.05. ROC curves have been performed on GraphPad Prism, and CI done with the Wilson-Brown Method. Optimal cutoffs were evaluated using the maximal Youden index. Logistic regression was performed using the “*fitglm*” function on MATLAB R2024a Update 6. Custom code used in this manuscript will be available at: https://github.com/LCIT-CHUV/Myocarditis

### Data Availability

Cytokines descriptive statistics data have been provided for the main figures. Corresponding FCS files and source data are available from the corresponding author, MO, upon reasonable request.

## Results

### Clinical and demographic characteristics of the cohort

A total of 33 cases of immune checkpoint inhibitor-related myocarditis (ICI-My) were identified at Lausanne University Hospital between July 2018 and June 2024, based on the 2022 European Society of Cardiology (ESC) diagnostic criteria as outlined in the Methods section. All 33 patients were included in the final analysis, with the study flowchart depicted in **Supplementary Figure 2**. Patients’ characteristics are summarized in **Table 1**. The mean age of the patients was 66.4 years, and there was a male predominance (25 out of 33 patients, 76%). The median time to ICI-My onset was 68 days (95% CI: 44–104 days). The median follow-up period following the initial ICI administration was 417 days (95% CI: 193–580 days). Lung cancer (13 out of 33 patients, 39%) and melanoma (8 out of 33 patients, 24%) were the most common cancer types among the cohort. The majority of patients received anti-PD-1 monotherapy (19 out of 33 patients, 58%) or a combination of anti-PD-1 and CTLA-4 inhibitors (9 out of 33 patients, 27%). The most frequent associated immune-related adverse events (irAEs) included colitis (24%), myocarditis-myositis overlap syndrome (21%), and endocrinopathies (4%). Regarding severity, 73% of patients developed low-grade myocarditis, while 27% experienced high-grade myocarditis. Arrhythmias at diagnosis were present in 57.5% of cases. Mean at diagnosis concentrations of cardiac biomarkers were as follows: cTnT at 359.4 ng/ml, cTnI at 159.6 ng/ml, and NT-proBNP at 3486 pg/ml. Additionally, 100% of patients had at least one elevated cardiac biomarkers (**Figure 1 and Supplementary Table 2**). In terms of treatment, 70% of patients were managed with corticosteroids (CS) including 24% received a combination of TCZ and CS, 12% a combination of infliximab and CS and 6% a combination of mycophenolic mofetil (MMF) and CS. Other characteristics, including imaging findings and electrocardiogram (ECG) data, are comprehensively detailed in **Supplementary Table 2.**

**Figure 1.**
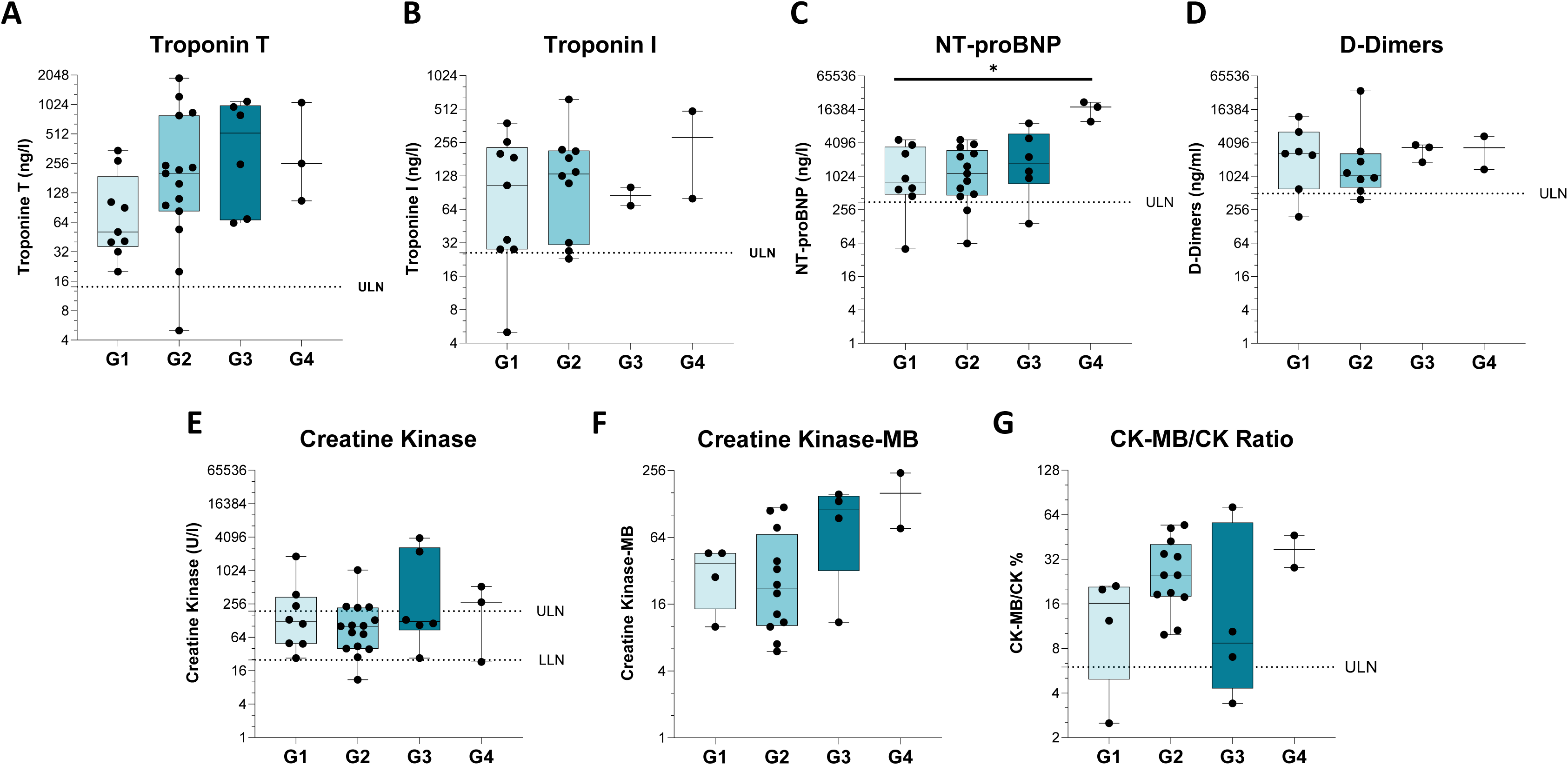
Biological profiles stratified by clinical grade of ICI-My. (A–G) Serum biomarker levels measured at the time of ICI-My diagnosis in patients: (A) serum cardiac troponin T (cTnT) (n=33), (b) serum cardiac troponin I (cTnI) (n=23), (C) NT-proBNP (n=30), (D) D-dimers (n=20), (E) Total creatine kinase (CK) (n=32), (F) Creatine kinase-MB (CK-MB) (n=22) and (G) CK-MB/CK ratio (n=22). Each plot displays individual patient values, with bars indicating mean levels and error bars representing standard deviations. Statistical analysis revealed significant differences between groups using Kruskal-Wallis Test (***P<0.001; ****P<0.0001). ULN denotes the upper limit of normal for each biomarker as defined by our institution.

**Table 1:** Patients, treatments, and clinical characteristics. The principal clinical, demographic, and therapeutic data for the entire cohort (n=33) are presented in detail in this table.

|  | N=33 | % within cohort |
| --- | --- | --- |
| **<u>Age</u>** |  |  |
| Median | 66.4 |  |
| ≥ 65 years | 15 | 45% |
| **<u>Gender</u>** |  |  |
| Female | 8 | 24% |
| Male | 25 | 76% |
| **<u>Time to myocarditis onset</u>** |  |  |
| From ICI treatment initiation (weeks, median, SD) | 9.7 (14.66) |  |
| **<u>Tumor type</u>** |  |  |
| Lung | 13 | 39% |
| Melanoma | 8 | 24% |
| ENT | 2 | 6% |
| Liver | 2 | 6% |
| Kidney | 2 | 6% |
| T cell lymphoma | 1 | 3% |
| Others | 5 | 15% |
| **<u>ICI treatment</u>** |  |  |
| Anti-PD1 | 19 | 58% |
| Anti-CTLA4/PD1 | 9 | 27% |
| Anti-PDL1 | 5 | 15% |
| **<u>Associated irAEs</u>** |  |  |
| Colitis | 8 | 24% |
| Myocarditis-myositis overlap syndrome | 7 | 21% |
| Hepatitis | 4 | 12% |
| Endocrinopathies | 4 | 12% |
| Pneumonitis | 3 | 9% |
| Arthralgia | 2 | 6% |
| Pancreatis | 2 | 6% |
| Peripheral neuropathy | 1 | 3% |
| Others | 10 | 30% |
| **<u>Myocarditis grading</u>** |  |  |
| Low Grade (G1-G2) | 24 | 73% |
| High Grade (G3-G4) | 9 | 27% |
| **<u>Biomarkers at diagnosis (mean (SD), n)</u>** |  |  |
| Troponine T | 359.4 (459.6, 33) |  |
| Troponine I | 159.6 (157.3, 23) |  |
| NT-proBNP | 3486 (5167, 30) |  |
| **<u>Immunosuppression therapy</u>** |  |  |
| Corticosteroids (CS) | 23 | 70% |
| Tocilizumab (TCZ) | 8 | 24% |
| Infliximab (IFX) | 4 | 12% |
| Intravenous immunoglobulin (IVIG) | 2 | 6% |
| Mycophenolic mofetil (MMF) | 2 | 6% |
| Colchicine | 1 | 3% |

### Comprehensive cytokine and biological profiles across different clinical grades of ICI-My

To characterize the inflammatory profiles of myocarditis, we analyzed an extensive panel of biological and inflammatory biomarkers in patients with varying clinical grades of immune checkpoint inhibitor-related myocarditis (ICI-My) (Grades 1 to 4). Cardiac biomarkers, including troponin T (cTnT), troponin I (cTnI), N-terminal pro-B-type natriuretic peptide (NT-proBNP), the CK-MB/CK ratio, and D-dimers, were elevated since grade 1 in all patients (**Figure 1**). NT-proBNP was shown to identify differences in NT-proBNP with grade 4 patient with increased levels of NT-proBNP (**Figure 1.C**). However, no others further significant differences in these markers were observed with increasing severity of myocarditis (**Figure 1**). Regarding cytokines, we observed substantial upregulation of IL-6, CXCL9, CXCL10, CXCL13, and sCD25, and a modest increase in CCL3, CCL4, CCL5, CXCL1, and VEGF-A. Notably, the distribution of CCL4 and CXCL12 differed between low (G1 and G2) and high-grade (G3 and G4) myocarditis with lower levels in high-grade cases (**Figure 2**). A correlation matrix analysis of biomarkers across all patients revealed several notable associations: CK-MB was positively correlated with troponin T, CK, and CXCL12. CXCL9, CXCL10, and CXCL13 each showed a positive correlation with sCD25. Additionally, CXCL9 was positively correlated with CXCL10. IL-6 showed a positive correlation with sCD25. BDNF was positively correlated with PDGF-BB (**Supplementary Figure 3**).

**Figure 2.**
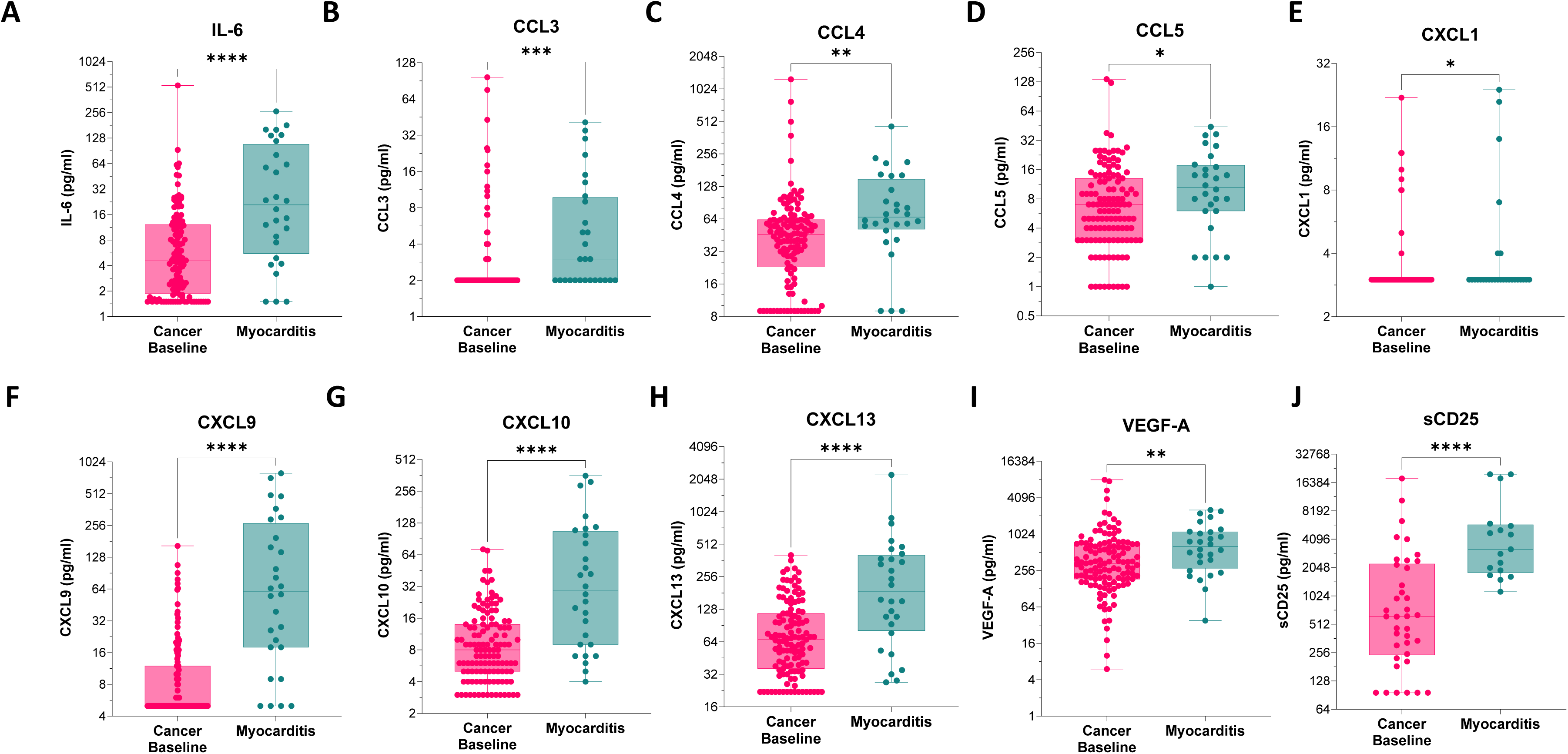
Comprehensive cytokine profiles stratified by clinical grade of ICI-My. (A–G) Serum levels of various chemokines, growth factors, and cytokines measured at ICI-My diagnosis in 28 patients with differing clinical grades, compared to pre-ICI serum levels obtained from 97 cancer patients serving as a reference group, baseline CD25 levels were available for 38 patients prior to ICI treatment and were compared to CD25 levels measured at ICI-My diagnosis in 17 patients. All comparisons were conducted using Mann-Whitney tests. Data are presented as individual values, with bars indicating the mean and error bars representing the standard deviation. Statistical significance was indicated at ***P < 0.001 and ****P < 0.0001.

The elevation of IL-6 and other inflammatory cytokines suggests a prominent role of the inflammatory response in ICI-My, which may inform targeted therapeutic strategies. The distinct cytokine profiles between low- and high-grade myocarditis highlight the potential for biomarker-based patient stratification, which could guide personalized treatment approaches and improve clinical outcomes. The observed correlations between biomarkers suggest potential interactions and common regulatory mechanisms that warrant further investigation. For example, the strong association between IL-6 and sCD25 may reflect coordinated immune activation, while the relationship between CXCL9, CXCL10 and sCD25 suggests a linked chemokine axis involved in immune cell recruitment and activation in myocarditis.

### The characterization of immune cell subsets distribution in ICI-My

Significant alterations within immune cell populations were observed among patients experiencing immune checkpoint inhibitor-related myocarditis (ICI-My). In particular, there was a notable reduction in mature neutrophils, accompanied by an increase in immature neutrophils **(Figure 3A**). Furthermore, there was a pronounced increase in activated double-positive HLA-DR/CD38+ T cells and a reduction in circulating CXCR5+ leukocytes **(Figure 3B-C**. This decline in CXCR5+ leukocytes was predominantly due to a reduction in both memory B cells and memory CD4+ T cells (**supplementary Figure 4A-D**). Moreover, CD4+CXCR5+ T cells displayed elevated levels of CD27, indicating their potential identity as peripheral T follicular helper (Tfh) cells (**supplementary Figure 4C**). Additionally, a notable decline in CXCR3 expression was observed across all CD8+ memory T-cell subsets (**Figure 3D-G**). A comparable, albeit less pronounced, reduction was observed in CD4+ memory T-cell subsets. It is noteworthy that there was no overall decrease in the frequency of circulating memory T cells (**supplementary Figure 4),** which suggests that the reduction in CXCR3+ T cells in patients with myocarditis is likely due to downregulation or internalization of CXCR3 rather than enhanced migration. This phenomenon may be driven by elevated circulating levels of CXCL9 and CXCL10.

**Figure 3.**
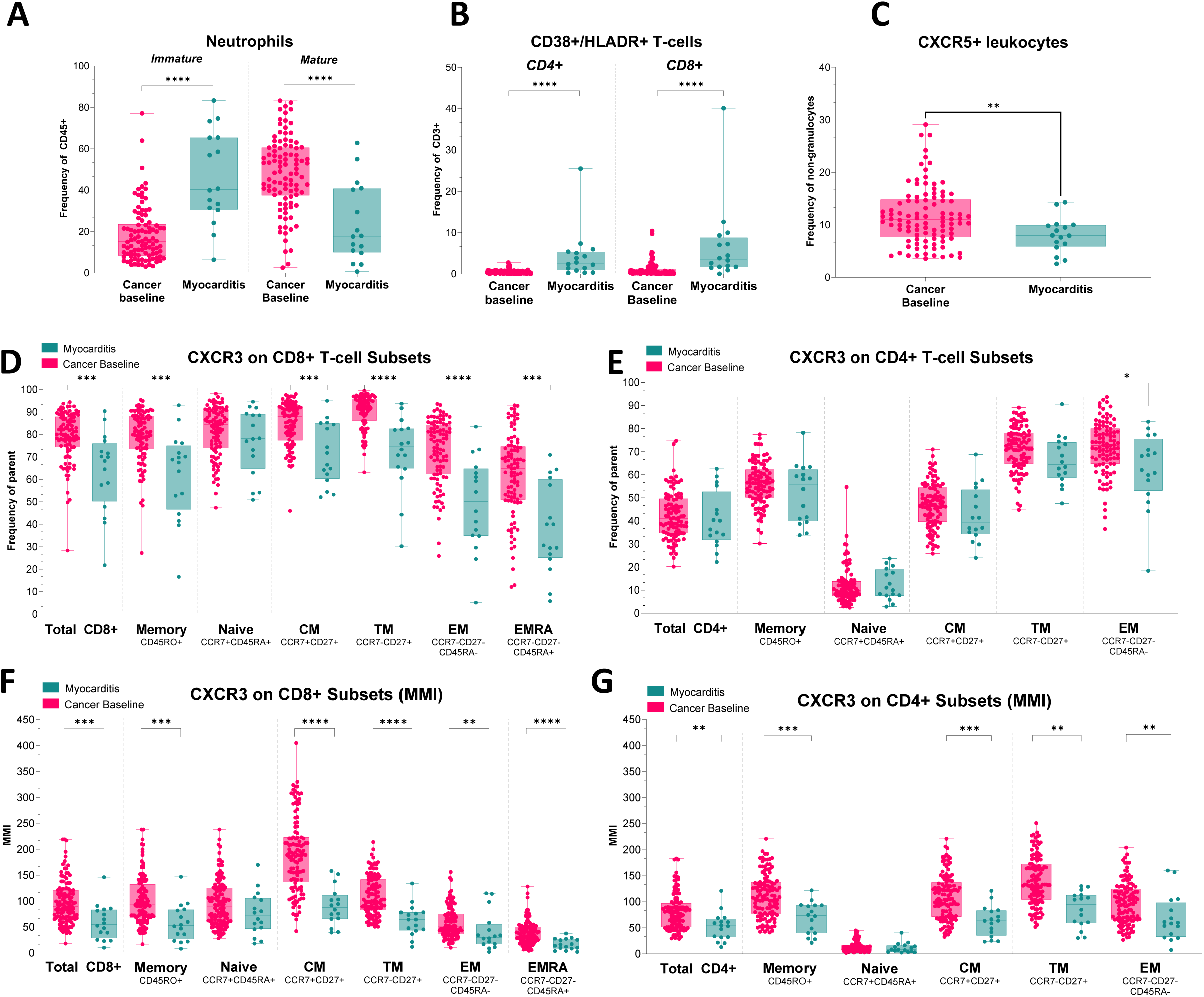
Distribution and phenotypes of circulating immune cell populations in myocarditis. (A) Frequencies of circulating immature (CD10–) and mature (CD10+) neutrophils expressed as a percentage of CD45+ cells, (B) Frequencies of activated T cells (CD38+HLA-DR+) within CD4+ and CD8+ T-cell subsets, expressed as a percentage of total CD3+ cells, (C) Frequency of CXCR5+ leukocytes expressed as a percentage of CD66b– cells, (D–G) Expression of CXCR3 on CD8+ and CD4+ T-cell subsets. (D–E) show the frequency of CXCR3+ cells, while (F–G) present the mean metal intensity (MMI) of CXCR3 expression. Data were obtained from 16 patients at ICI-My diagnosis and compared with baseline levels from 98 cancer patients. Statistical significance was determined using the Mann-Whitney U-test, with *P<0.05, **P<0.01, ***P<0.001, and ****P<0.0001.

The phenotype of circulating B cells was markedly altered in patients with myocarditis, exhibiting indications of activation as evidenced by a reduction in the expression of CD19, CD45RA, and IgM. However, no increase in other immunoglobulin isotypes was observed, suggesting that class-switch recombination may be impaired (**supplementary Figure 4F**). This impairment may be attributed to the deficiency of peripheral Tfh cells (pTfh), and the disruption of chemotactic signals caused by elevated levels of circulating chemokines (**supplementary Figure 4**). The observed shifts in neutrophil maturity suggest an acute inflammatory response, while the increase in double-positive HLA-DR/CD38 T cells indicates heightened T cell activation. The reduction in CXCR5+ leukocytes, particularly memory B-cells and memory CD4+ T-cells, alongside decreased CXCR3 expression, suggests the potential for dysregulation in immune surveillance and response mechanisms. The observed impairment in class-switch recombination within B-cells, as evidenced by reduced CD19, CD45RA, and IgM levels without corresponding increases in other immunoglobulin isotypes, may hinder effective antibody-mediated responses. This could contribute to the persistence or exacerbation of inflammatory processes within the myocardium. Furthermore, no notable differences were observed in the frequencies of the three monocyte subsets (classical, intermediate, and non-classical) or in their mean metal intensity (MMI) for various activation and functional markers (CD11c, HLA-DR, CD141, CD31, CD38, CD123, CD62L, CD69, and CD1c). Similarly, the expression levels of chemokine receptors (CCR4, CCR6, CCR7, CXCR3, and CXCR5) were found to be comparable across the various subsets **(supplementary Figure 4)**.

### Evaluation of complement system involvement in ICI-induced myocarditis

To investigate the potential involvement of the complement system in ICI-My, we conducted a comprehensive assessment of the different complement activation pathways. Specifically, we evaluated components from the classical, alternative, and terminal common pathways. The classical pathway was assessed by measuring C4 and CH50 levels, the alternative pathway was evaluated by measuring Factor Bb and C3c levels and the terminal pathway was assessed by measuring of the terminal complement complex C5-b9 levels, representing the formation of the membrane attack complex (MAC). Our analysis revealed no evidence of activation in any of the assessed complement pathways among ICI-My patients. Specifically, levels of C4, C3c, CH50, Factor Bb, and C5-b9 remained within the normal range across (**Figure 4**). This absence of complement activation suggests that the complement system may not play a significant role in the pathogenesis of ICI-My, or that its activation is not a prominent feature in this condition.

**Figure 4.**
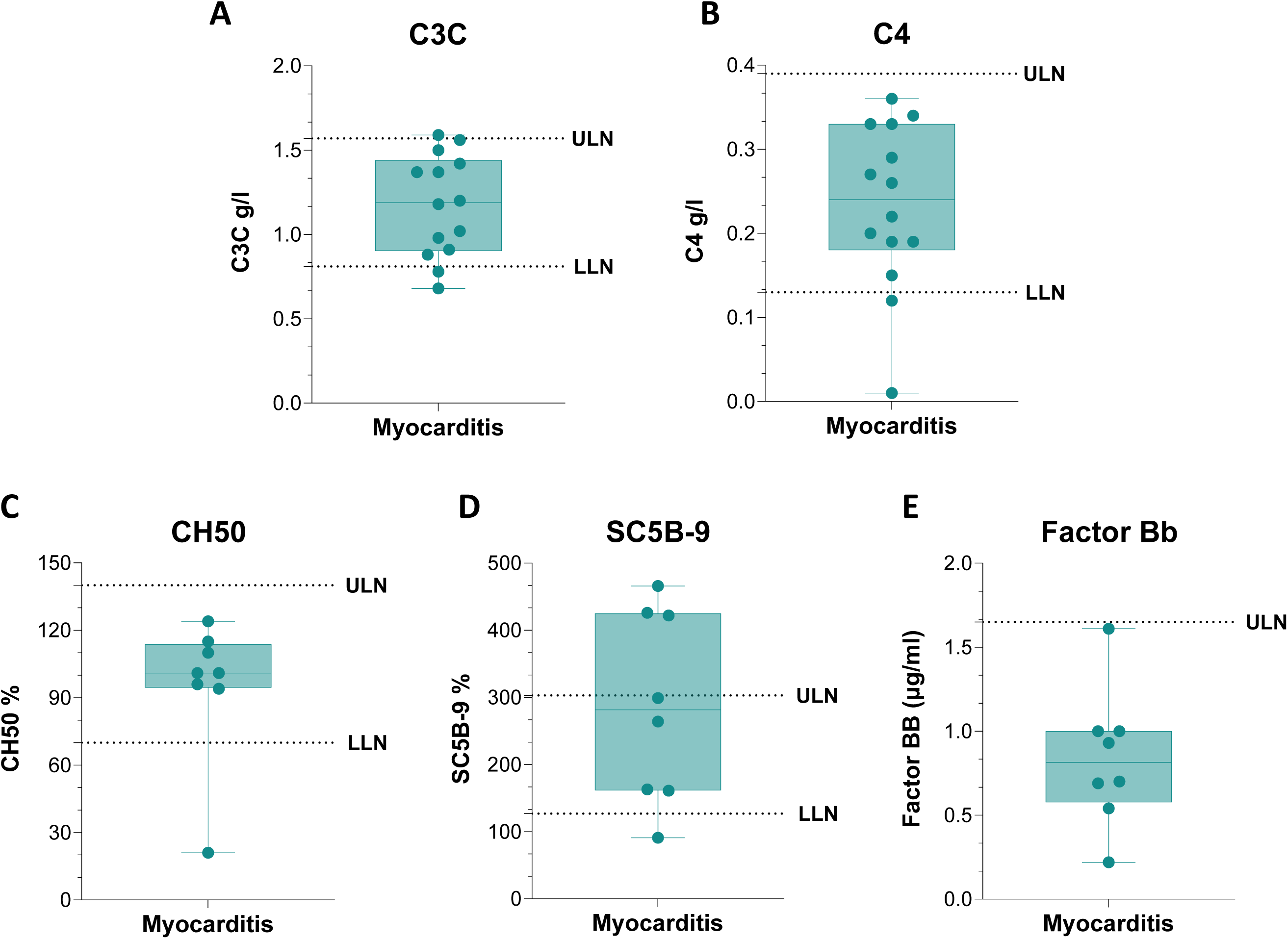
Evaluation of complement system involvement in ICI-My. Levels of key components representing the classical (C4 n=14, CH50 n=8), alternative (Factor Bb n=8, C3c n=14), and terminal (C5-b9 n=8) complement activation pathways were measured at ICI-My diagnosis. Statistical significance was determined using the Mann-Whitney U-test, with *P<0.05, **P<0.01, ***P<0.001, and ****P<0.0001.

### Distinct cytokine signatures differentiate low- and high-grade myocarditis

In order to investigate the relationship between cytokine levels and clinical severity, we conducted a comparative analysis of the concentrations of all measured biomarkers and cytokines in patients with low-grade versus high-grade immune checkpoint inhibitor-related myocarditis (ICI-My). NT-proBNP and CK-MB were found to be increased in High-grade compared to low grade myocarditis **(Figure 5A).** Furthermore, both showed good performance to discriminate Low and High grades ICI-my. NT-proBNP cutoff value of 4770 ng/l showed a 55.56% Sensitivity (95% CI 26.7% to 81.1%) but a 100% Specificity (95% CI: 56.99% to 93.41%), CK-MB cutoff value of 61.5 U/l showed a 83.3% Specificity (95% CI: 43.65% to 99.15%) and a 81.25% Specificity (95% CI: 56.99% to 93.41%) **(Supplementary Figure 5)**. Using optimal cutoff, NT-proBNP showed 100% PPV (predicting low grades) and CK-MB 93% NPV (predicting high grades) **(Supplementary Figure 6C** to E). Others traditional cardiac biomarkers troponin I, D-dimers, CK, CK-MB/CK ratio did not yield statistically significant results in differentiating between low- and high-grade disease severity (**Figure 5A**), clustering approaches also failed with ROC AUC curve ranging from 0.51 to 0.71 and p value from 0.93 to 0.08 (**Data not shown)**. Among the evaluated cytokines, only CCL4 and CXCL12 exhibited significant differences between the two groups, with both cytokines displaying lower levels in the group with high-grade myocarditis (**Figure 5B**). Specifically, a CCL4 concentration below 59.5 pg/mL was found to be an effective discriminant between high-grade and low-grade disease, demonstrating a sensitivity of 85.71% (95% CI: 48.69%–99.27%) and a specificity of 76.19% (95% CI: 54.91%–89.37%). Similarly, a CXCL12 concentration below 613.5 pg/mL yielded a sensitivity of 85.71% (95% CI: 48.69%–99.27%) and a specificity of 80.95% (95% CI: 60%–92.33%) for distinguishing high-grade myocarditis **(Supplementary Figure 5).**

**Figure 5.**
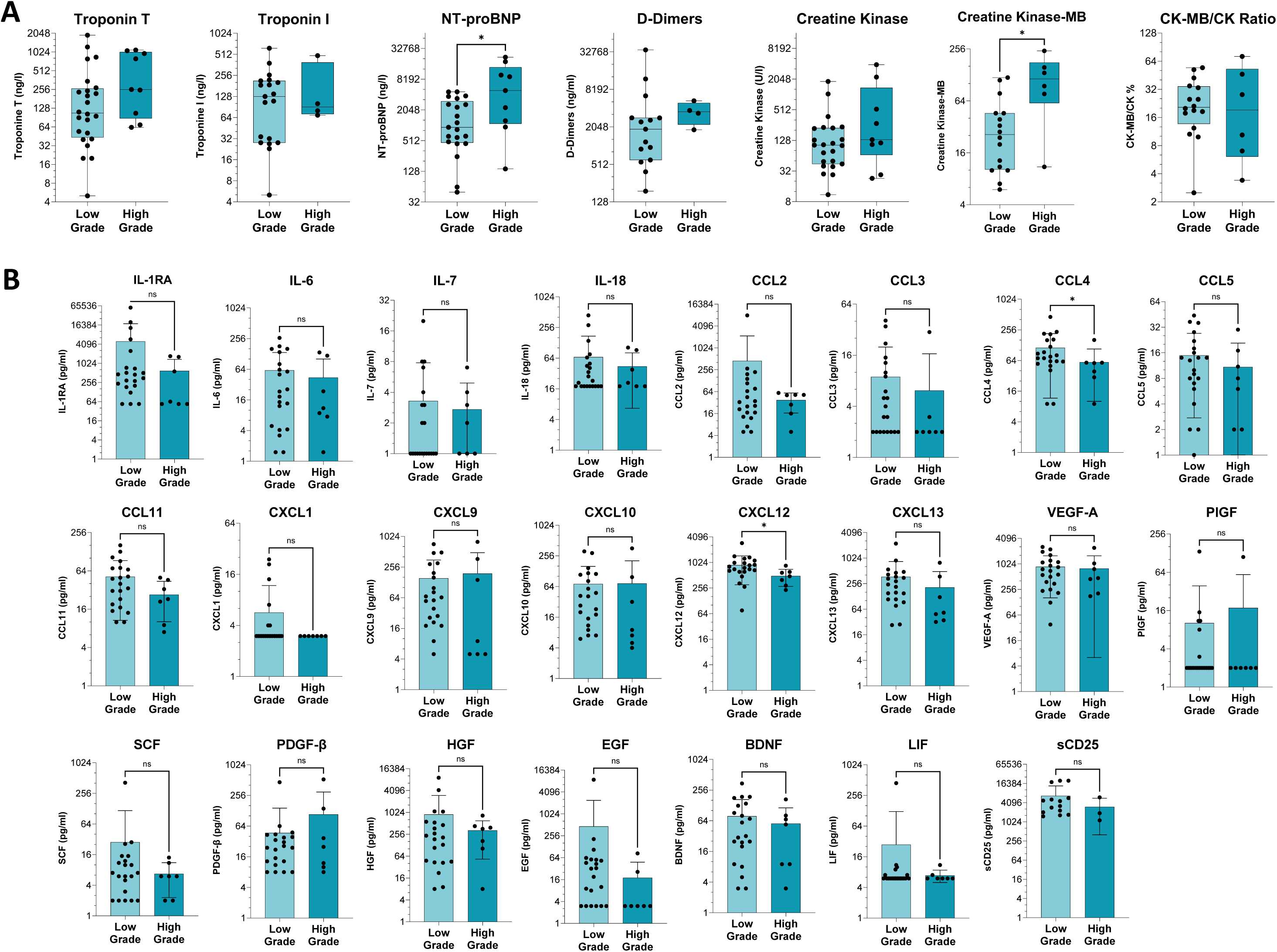
Distinct cytokine signatures differentiate low- and high-grade ICI-My. (A) Comparative analysis of classical biomarkers comparing serum cardiac troponin T (cTnT) (n=33), serum cardiac troponin I (cTnI) (n=23), NT-proBNP (n=30), D-dimers (n=20) Total creatine kinase (CK) (n=32), Creatine kinase-MB (CK-MB) (n=22) and CK-MB/CK ratio (n=22) in high and low grade ICI-My diagnosis. (B) Serum chemokines, growth factors, and cytokines at ICI-My diagnosis, examining 21 patients with low-grade and 7 patients with high-grade disease. Statistical significance was determined using the Mann-Whitney U-test, with *P<0.05, **P<0.01, ***P<0.001, and ****P<0.0001). ULN denotes the upper limit of normal for each biomarker as defined by our institution.

Regarding immune subset differences, no major differences were observed between low- and high-grade myocarditis. However, given the limited number of high-grade patients analyzed, these results should be interpreted with caution (**Supplementary Figure 6**). The marked reduction in CCL4 and CXCL12 levels in high-grade myocarditis suggests their potential as novel biomarkers for accurately assessing disease severity in ICI-My. In contrast to traditional cardiac markers, these cytokines demonstrate superior diagnostic performance, which could facilitate earlier identification and more tailored therapeutic interventions for patients at risk of severe myocarditis.

### Tocilizumab as an effective treatment for corticosteroid (CS)-refractory ICI-My

Given the marked IL-6 upregulation observed in ICI-related myocarditis (ICI-My) and our prior report of a corticosteroid (CS)-refractory ICI-My case successfully managed with TCZ^19^, we expanded our investigation to a larger cohort of CS-refractory ICI-My patients including seven (n=8) patients. This study was conducted with a particular focus on patients who had not responded to other immunosuppressive treatments, including mycophenolate mofetil (MMF), and anti-TNFα therapy. The clinical characteristics of the cohort are presented in **Supplementary Table 3.** Melanoma (37%) and Lung cancer (24%) were the most prevalent tumor types, with 75% of patients receiving combination anti-CTLA-4/anti-PD-1 regimens and 25% receiving anti-PD-1 therapy alone. Colitis was the most common concurrent immune-related adverse event (irAE), affecting 50% of cases. At the time of initial presentation, 38% of patients exhibited high-grade myocarditis. A subset of seven (n=7) patients demonstrated refractory myocarditis despite the administration of high-dose corticosteroids (HDS) (dosages ≥ 1 mg/kg). Of these patients, two received infliximab, one received mycophenolate mofetil (MMF), one received intravenous immunoglobulin (IVIG), one received colchicine, and one received rituximab. The therapeutic response to TCZ treatment was favorable in all patients, with no recorded fatalities related to myocarditis. Furthermore, successful tapering of corticosteroids was achieved by all patients. It is important to note that 50% of patients required only a single TCZ dose. In addition, the initiation of TCZ resulted in a rapid decrease in IL-6, CXCL1, CXCL9, CXCL10, CXCL13, sCD25, CCL3, CCL4, CCL5, and VEGF-A, as well as in cardiac biomarkers (troponin T and NT-proBNP) (**Figure 6**). The kinetics of biomarker reduction were comparable to those observed in patients treated solely with other immunosuppressive agents (**Supplementary Figure 7**).

**Figure 6.**
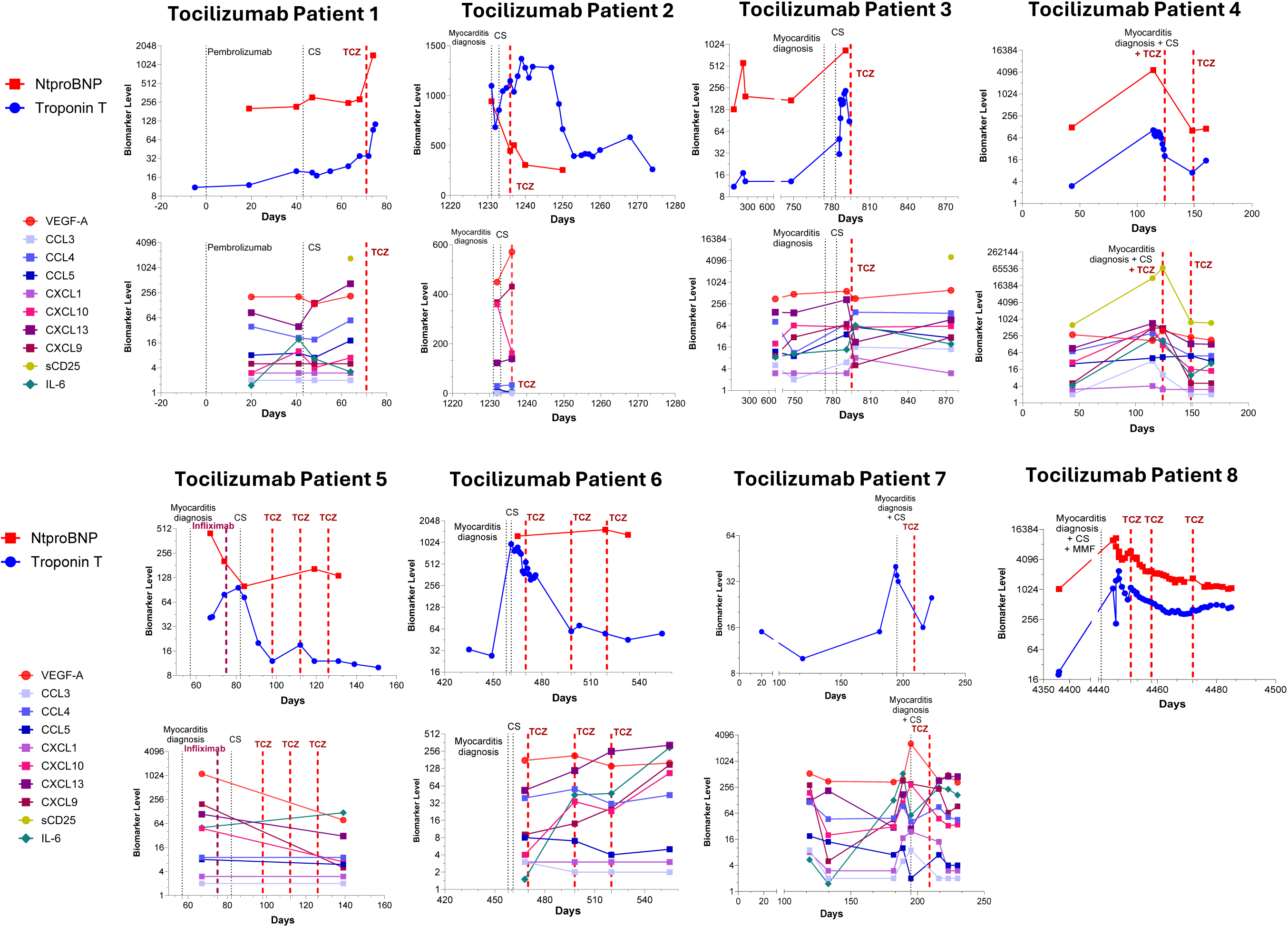
Longitudinal biomarker changes and evidence of biological resolution in corticosteroid (CS)-refractory ICI-Myocarditis patients treated with tocilizumab (TCZ). Each panel represents an individual patient (n=8) treated with TCZ after inadequate response to CS therapy. Longitudinal changes in key cardiac biomarkers (cTnT [ng/L], NT-proBNP [ng/L]) and accompanying dynamics in selected cytokines, chemokines, and growth factors are shown over time. The onset date of myocarditis and the timing of immunosuppressive treatments are indicated on each patient’s panel. Abbreviations: CS, corticosteroids; TCZ, tocilizumab.

In summary, the present findings demonstrate that TCZ is a highly effective therapy for CS-refractory ICI-My, including cases unresponsive to multiple immunosuppressive agents. The rapid clinical and biological improvement observed in all treated patients, concomitant with the absence of related mortality, supports the addition of anti-IL-6R therapy as a potentially life-saving treatment option for severe ICI-My, with the potential to improve patient outcomes and mitigate life-threatening complications.

### Cytokine performance in predicting the need for immunosuppressive treatment and tocilizumab intensification

In this study, we investigated the predictive performance of cytokines and biological biomarkers in determining the necessity for immunosuppressive (IS) therapy and the intensification of treatment with TCZ in patients with ICI-Myo. Specifically, optimal cutoff values were identified for IL-18 (>31 pg/ml), CCL2 (>48 pg/ml), BDNF (>24.5 pg/ml), and HGF (>105.5 pg/ml), which significantly predicted the need for IS therapy in the myocarditis group treated with IS agents, including tocilizumab, compared to those who did not require IS treatment. These cutoffs achieved accuracies ranging from 71% to 89% in discerning individuals who required IS therapy (**Figure 7A-7B**). Moreover, we assessed the performance of select cytokines in predicting the specific need for TCZ within this cohort. An IL-18 level exceeding 36 pg/ml is a highly accurate predictor of TCZ treatment, with a sensitivity of 85.71% (95% CI: 48.69%–99.27%) and a specificity of 76.19% (95% CI: 54.91%–89.37%). Similarly, a CCL4 level below 56 pg/ml was identified as a significant predictor for the need for TCZ treatment, demonstrating a sensitivity of 71.43% (95% CI: 35.89%–94.92%) and a specificity of 85.71% (95% CI: 65.36%–95.02%) (**Figure 7B**). The identification of these specific cytokine cutoffs—particularly for IL-18 and CCL4—provides valuable biomarkers for predicting the necessity of immunosuppressive therapy and potential escalation to TCZ in patients with ICI-Myo. Further analysis using multivariate logistic regression, while limited in this context, improved accuracy to 93.75%, highlighting the potential of combining multiple biomarkers (here IL-7,-18, CXCL10, CXCL13 and BDNF) to enhance the prediction of treatment needs **(Supplementary Figure 8)**. Additionally, classical biomarkers showed no significant differences between patients who received immunosuppressive treatment and to who did not received treatment (**Supplementary Figure 9).** By demonstrating superior predictive power relative to conventional cardiac markers, these cytokines enable more precise, personalized treatment strategies aimed at improving patient outcomes and reducing mortality in refractory myocarditis cases.

**Figure 7.**
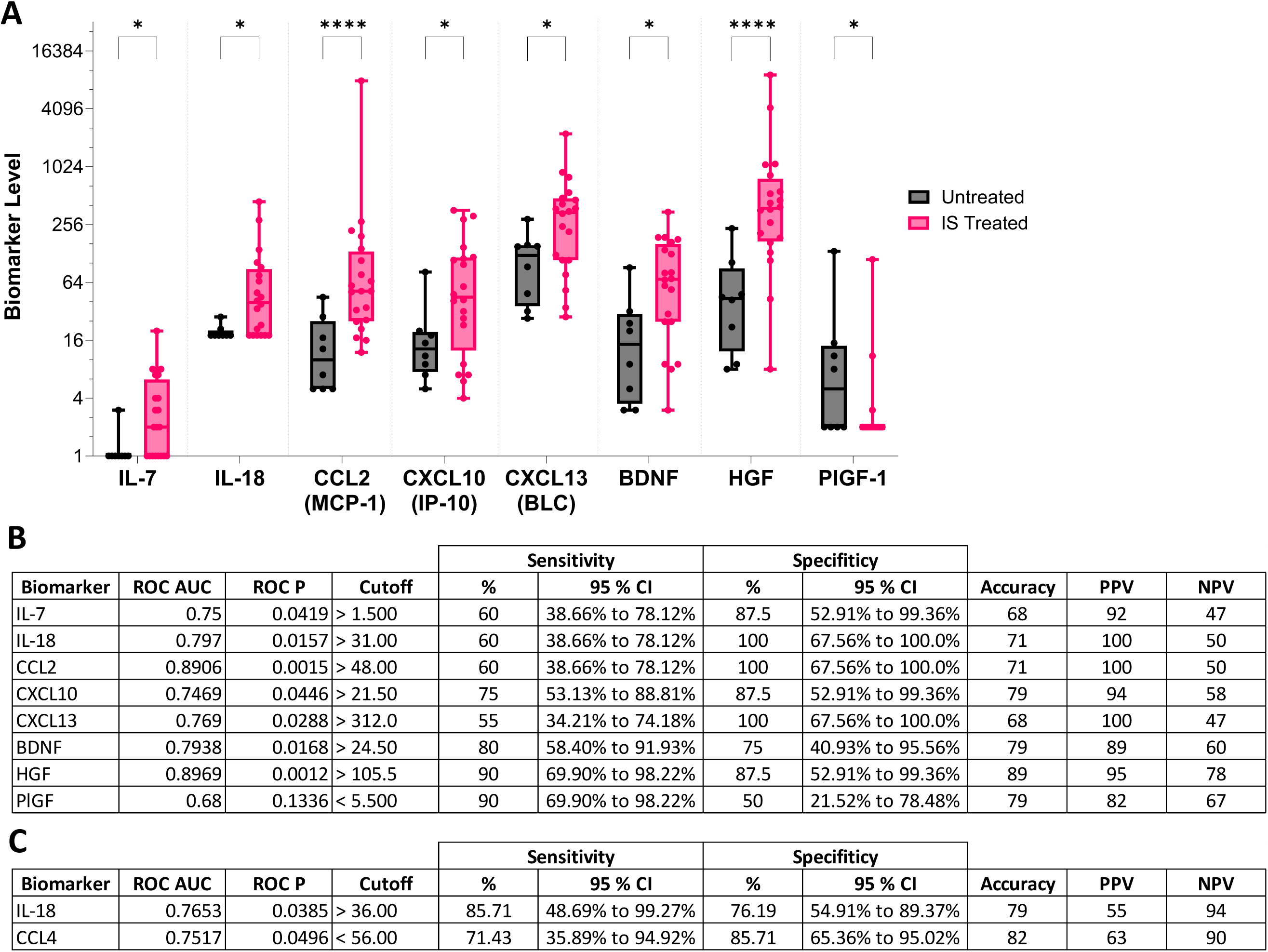
Cytokine performance in predicting the need for immunosuppressive (IS) therapy and tocilizumab (TCZ) intensification in ICI-induced myocarditis (ICI-My). (A) Comparison of cytokine levels at ICI-My diagnosis between patients who received IS treatment (including TCZ) (n=20) and those who did not (n=8), (B) Identification of optimal cytokine cutoff values—IL-18 (>31 pg/mL), CCL2 (>48 pg/mL), BDNF (>24.5 pg/mL), and HGF (>105.5 pg/mL)—that predict the absence of IS treatment requirements, (C) Within the TCZ-treated cohort (n=7), IL-18 (>36 pg/mL) and CCL4 (<56 pg/mL) emerged as accurate predictors for the need to intensify TCZ therapy compared to all other patients (n=21). These cytokine-driven thresholds offer superior predictive accuracy over conventional cardiac biomarkers, guiding more personalized and effective therapeutic decision-making. Such stratification may improve patient outcomes and reduce mortality in refractory ICI-My cases.

## Discussion

This study provides a comprehensive immunological characterization of ICI-My, combining an extensive cytokine panel and advanced mass cytometry profiling with traditional cardiac biomarkers and imaging assessments. By elucidating the immune landscape that underlies ICI-My inflammation, this integrated approach offers new insights into the immunopathogenesis of ICI-My and highlights potential biomarkers for improved diagnosis, risk stratification, and treatment optimization. The principal objective of this study was to generate hypotheses rather than to draw definitive conclusions.

Patients with ICI-My were characterized by significant increases in several cytokines, including IL-6, CXCL9, CXCL10, CXCL13, VEGF-A, and sCD25, as well as intermediate elevations in CCL3, CCL4, CCL5, and CXCL1. These findings underscore the presence of a robust cytokine-mediated response and suggest the existence of a dynamic interplay between inflammatory, chemotactic, and tissue remodeling factors within the myocardium. IL-6, a pro-inflammatory cytokine, has been implicated in numerous autoimmune and inflammatory processes^29^, as well as in various irAEs^19,26–28,30,31^. The pronounced elevations in IFNγ-inducible chemokines CXCL9 and CXCL10 may indicate an intensified trafficking of T cells to the inflamed myocardial tissue^32^. The increased expression of CXCL13, an important regulator of B-cell localization, suggests a potential role for B cells in the pathogenesis of ICI-related myocarditis^33^. Elevated VEGF-A may indicate endothelial activation or compensatory angiogenesis^34^, while sCD25, a marker of T-cell activation, provides further evidence for the hypothesis that an active, T-cell-driven inflammatory milieu is present^35^. The intermediate increases in CCL3, CCL4, CCL5, and CXCL1 suggest the presence of a complex chemokine network that is responsible for the recruitment and activation of diverse immune cell subsets, including monocytes, T cells, B cells and neutrophils^36^. In contrast with certain inflammatory or autoimmune conditions where complement activation represents a central phenomenon, our study did not detect complement pathway activation in ICI-related myocarditis. The presence of normal levels of C4, C3C, CH50, SC5B-9, and factor Bb indicates that complement involvement is minimal or non-essential in this context. This finding aligns with growing evidence that cytokines and T-cell– mediated mechanisms are the principal drivers of ICI-related irAEs^36^. The lack of complement activation may have therapeutic implications, focusing attention on cytokine-mediated and cellular immune processes rather than complement inhibition.

It is clinically relevant to note that the levels of CCL4 and CXCL12 differentiate high-grade and low-grade ICI-induced myocarditis. The lower levels of these cytokines observed in high-grade cases, with robust ROC performance (AUC values of 0.75 and 0.79), highlight their potential as biomarkers of disease severity. CXCL12 is recognized for its function in the homing of stem cells and the repair of tissues, as well as in the context of various cardiovascular diseases^37^. A reduction in CXCL12 in severe disease may indicate impaired regenerative responses or excessive tissue damage. CCL4, which plays a pivotal role in the recruitment of regulatory T cells^38^, was also diminished in high-grade cases, suggesting a disrupted chemokine milieu in more severe myocarditis. NT-proBNP and CK-MB increased in high-grade myocarditis confirm that more severe myocardial damage is to be found in High Grade patients. Furthermore, only CK-MB and not CK was found in increased in high-grade myocarditis confirming that the increased grading is associated with myocardial damage, more than global muscular damage. In contrast, traditional biomarkers, including troponin I, troponin T, CK, were unable to differentiate between low and high-grade myocarditis. This underscores the greater discriminative capacity of cytokine profiling in comparison to conventional biomarkers for the assessment of disease severity and acknowledging the need for prompt and efficient intervention in High grade patients. The incorporation of cytokine levels into clinical practice could facilitate enhanced risk stratification, thereby enabling the implementation of tailored therapeutic interventions based on immunological profiles.

Additionally, it is possible that other types of severe ICI-related myocarditis may exhibit distinct immune profiles. This variability underscores the importance of thoroughly characterizing the various forms of severe ICI-My within a larger cohort where the performance of traditional biological markers could be deeply explored compared to cytokine. By delineating whether different immune signatures exist among severe ICI-My subtypes, we can potentially refine diagnostic accuracy and improve the utility of conventional biomarkers in assessing disease severity and guiding treatment strategies.

These findings underscore the potential of specific cytokines—including IL-18, CCL2, BDNF, HGF, and CCL4—to serve as valuable biomarkers for guiding therapeutic decisions in ICI-My. While traditional cardiac biomarkers often assess cardiac injury and inflammation, their prognostic and predictive utility for IS treatment requirements remains limited. In contrast, the cytokine cutoffs identified here not only demonstrated a strong association with the need for IS therapy but also offered actionable guidance on when to intensify immunosuppression with TCZ. Stratifying patients based on cytokine levels has several implications for clinical practice. First, it may enable earlier and more precise intervention. Patients exceeding these thresholds could be promptly considered for IS therapy or TCZ intensification, possibly limiting the progression of myocarditis and improving outcomes. Second, the use of cytokines as predictive biomarkers represents a more biologically grounded approach. Because cytokines directly mediate immune-driven inflammation, their concentrations likely reflect underlying pathophysiological processes more accurately than do generic cardiac injury markers.

By identifying IL-18 and CCL4 as especially predictive of the need for TCZ escalation, clinicians can determine which individuals fail to respond adequately to corticosteroids or other immunosuppressive agents and thus adapt treatment regimens accordingly. Moving toward a biomarker-driven approach may reduce unnecessary exposure to broad-spectrum immunosuppression and its associated risks, while ensuring that those at higher risk receive the intensified therapy they require. In conclusion, these results provide a foundation for more personalized management strategies in ICI-My, with the potential to significantly enhance patient outcomes and reduce mortality in refractory cases.

The results of mass cytometry analyses demonstrated pronounced alterations in circulating immune cells, thereby elucidating potential cellular mechanisms underlying ICI-related myocarditis. It is conceivable that the elevation of immature neutrophils may be indicative of an acute and ongoing inflammatory process with the potential to damage myocardial tissue. This finding is consistent with previous reports on the pathogenic implications of neutrophils in multiple cardiovascular inflammatory diseases^39,40^. Additionally, a reduction in memory B cells was observed in both switched and unswitched memory B cells, which may indicate an aberrant humoral response or selective trafficking of these cells into the inflamed myocardium^41^. The presence of activated T cells, as indicated by an increased number of double-positive CD38/HLA-DR T cells among both CD4+ and CD8+ subsets, suggests that T cells are actively involved in tissue injury^42^. A reduction in CXCR5+ leukocytes was observed. The diminished levels of CXCR5+ memory B and T cells in the circulation suggest the existence of active follicular trafficking and local inflammation in the myocardium^43,44^.

The reduction in CXCR3 expression on T cells, despite elevated CXCL9/10, suggests either intense chemokine-driven recruitment into inflamed tissue or receptor modulation in response to chronic stimulation^45^. Our findings support a model in which elevated chemokines (CXCL9, CXCL10, CXCL13) serve as drivers of immune cell migration into the myocardium. This trafficking diminishes the peripheral pool of receptor-expressing cells (CXCR3⁺, CXCR5⁺) and may contribute to disease progression or severity through localized immune activation and tissue damage. Similarly, the interplay between increased IL-6 and aberrant T- and B-cell dynamics underscores the key role of IL-6 as a central mediator within the inflammatory cascade, influencing both cellular activation and recruitment^29^.

Our findings suggest that anti-IL-6 therapy may be a promising intervention for patients with refractory ICI-My who do not respond to multiple lines of immunosuppression, including MMF and infliximab (anti-TNFα). This evidence highlights IL-6 as a pivotal mediator of the pathological immune response and establishes IL-6 blockade as a promising novel therapeutic strategy for cases that are refractory to conventional management. Moreover, the integration of anti-IL-6 therapy into existing treatment guidelines, as exemplified by the ESMO guidelines^18^, could facilitate a more precise and tiered approach to the management of ICI-induced myocarditis. The early identification of patients at high risk for severe disease or poor response to standard immunosuppressants could facilitate the prompt initiation of IL-6 blockade, which may improve outcomes and reduce both the duration and intensity of corticosteroid use. One of the most significant challenges in immunotherapy is achieving an optimal balance between antitumor efficacy and immune-related toxicity. If anti-IL-6 therapy is shown to be an effective means of mitigating severe myocarditis, it may permit patients to continue or resume potentially life-saving ICIs without compromising their cardiac health^28^. This paves the way for more flexible and durable treatment regimens, thereby improving long-term disease control.

As biomarker research advances, combining cytokine profiling with routine cardiac monitoring may help identify patients most likely to benefit from early IL-6 blockade. Stratifying patients based on IL-6 or related cytokine levels could enable clinicians to tailor therapy to individual immune and inflammatory profiles, thus minimizing trial-and-error approaches and enhancing cost-effectiveness. It is possible that IL-6 inhibition may have broader applications in other severe irAEs beyond myocarditis as we reported before in cholangio-hepatitis^27^, myocarditis^19^, CRS^46^, irHLH^26,46^, esophageal stenosis^30^ and arthritis^47^. Further research may investigate whether the combination of anti-IL-6 agents with ICIs, either preemptively or at the earliest indications of toxicity, could enhance the therapeutic window of checkpoint inhibition^28^. Such a strategy may allow for more aggressive or prolonged ICI dosing schedules, potentially enhancing tumor control while minimizing immune-mediated damage to the heart and other organs. While the potential of IL-6 blockade is promising, further investigation is required to address several limitations. The efficacy of anti-IL-6 therapy may be contingent upon the specific tumor type, ICI regimen, and the presence of preexisting comorbidities.

Further research is required in the form of larger, prospective, multicenter trials in order to establish efficacy, optimal dosing, and patient selection criteria. The long-term efficacy of anti-IL-6 therapy remains uncertain. Longitudinal follow-up is essential to ascertain whether IL-6 blockade results in sustained improvements in cardiac function, quality of life, and overall survival, as well as to monitor potential adverse effects associated with long-term immunomodulation. The large validation cohort may lead to the integration of cytokine thresholds into clinical decision-making, enabling clinicians to tailor interventions to the patient’s underlying immune dysregulation, with the potential to improve outcomes and reduce unnecessary exposure to broad-spectrum immunosuppression. The comparison with a large baseline cancer cohort (n=97) provides robust reference values, thereby enhancing the reliability of our conclusions^31^. This integrative approach—combining cytokines, traditional biomarkers, and mass cytometry—provides a comprehensive and multifaceted understanding of ICI-related myocarditis and its underlying immunopathology.

Despite the advances that have been made in our understanding of ICI-related myocarditis, several limitations remain. Further research is required in the form of larger, multicenter studies to confirm the generalizability of these findings. Future research should validate key biomarkers (e.g., CCL4, CXCL12) in independent cohorts, examine time-course changes, and evaluate targeted immunotherapies (e.g., anti–IL-6) in controlled clinical trials. In addition, further investigation into neutrophil and B-cell functions, the roles of CXCR5 and CXCR3, and the influence of T- and B-cell interactions is recommended. This multidimensional approach may facilitate novel therapeutic target discovery and the development of more precise risk stratification strategies, ultimately improving patient outcomes.

In conclusion, this study elucidates the intricate immune landscape of ICI-induced myocarditis, identifies cytokine biomarkers that surpass traditional markers in discriminating disease severity, and demonstrates the potential of anti-IL-6 therapy in refractory cases. By integrating cytokine profiling and mass cytometry findings, we present a more detailed and biologically informed framework for understanding, diagnosing, and managing ICI-related myocarditis. This framework promises to drive the development of personalized, biomarker-driven therapies.

## Acknowledgements

We thank the patients and their families. This work was supported by the strategic plan of the CHUV. We would like to express our gratitude to all the patients who generously contributed their time and samples for this project. Visual abstract was created with BioRender.com.

## Author contributions

M.O. and S.B., D.D. had full access to all the data in the study and take responsibility for its integrity and accuracy. MO conceived and designed the study and drafted the manuscript. S.B., D.D. R.B. and M.O. analyzed and interpreted the data. D.D., S.B., and M.O. collected the data. SB, SL, HB, JD, NF, KA, NM, LM, NE, KZ, KS, and SP participated in the clinical treatments. V.J., R.B. and AN analyzed the CyTOF data. D.D. SB, RB and M.O. prepared the figures. MO and RB prepared the visual summary. MO, MP, CF, GP supervised the panel development. JM and JEM participated in the scientific discussion. The manuscript was reviewed and approved by all authors before submission.

---

**Supplementary Figure 1:**
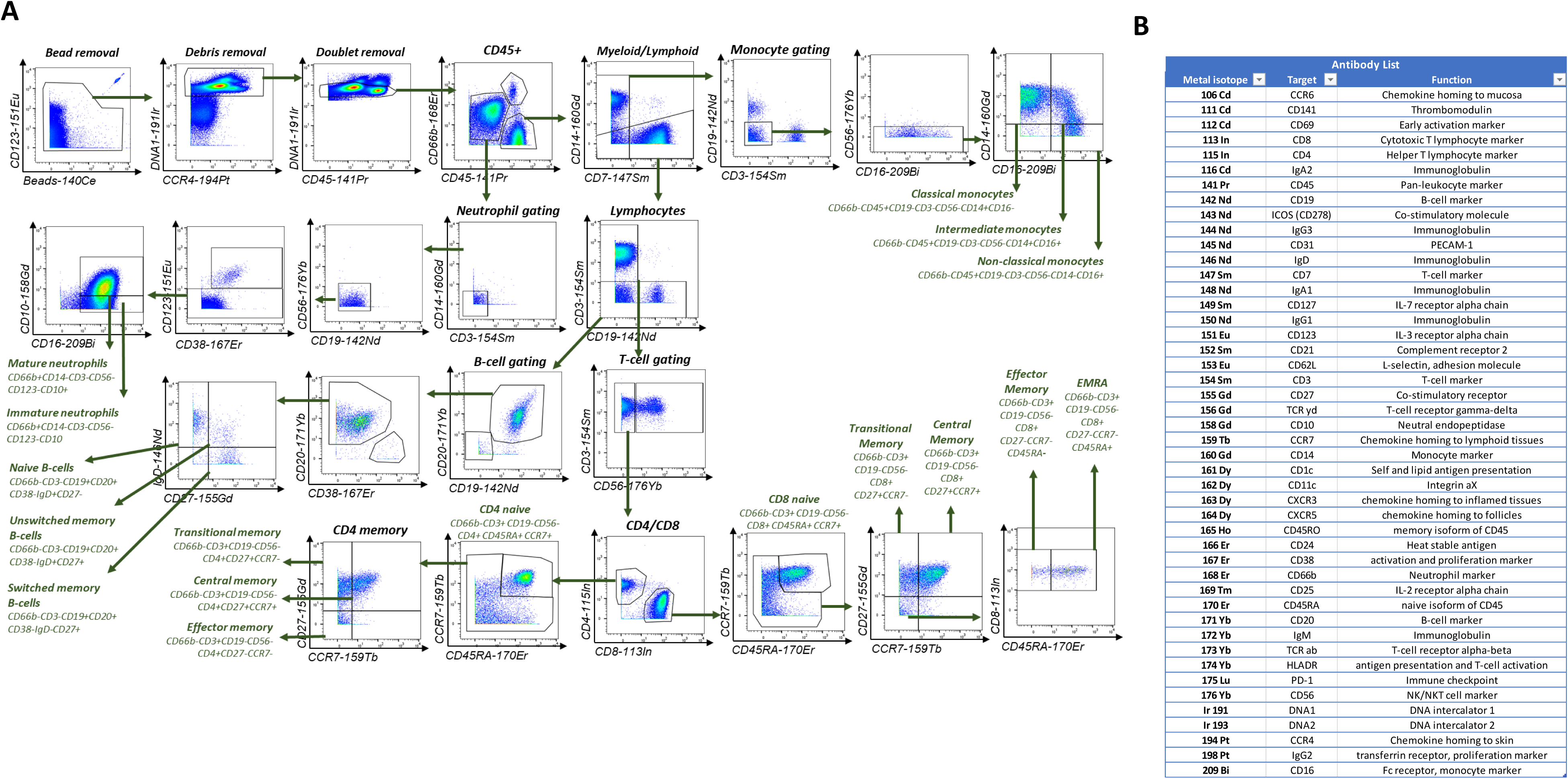
Study flowchart.

**Supplementary Figure 2:**
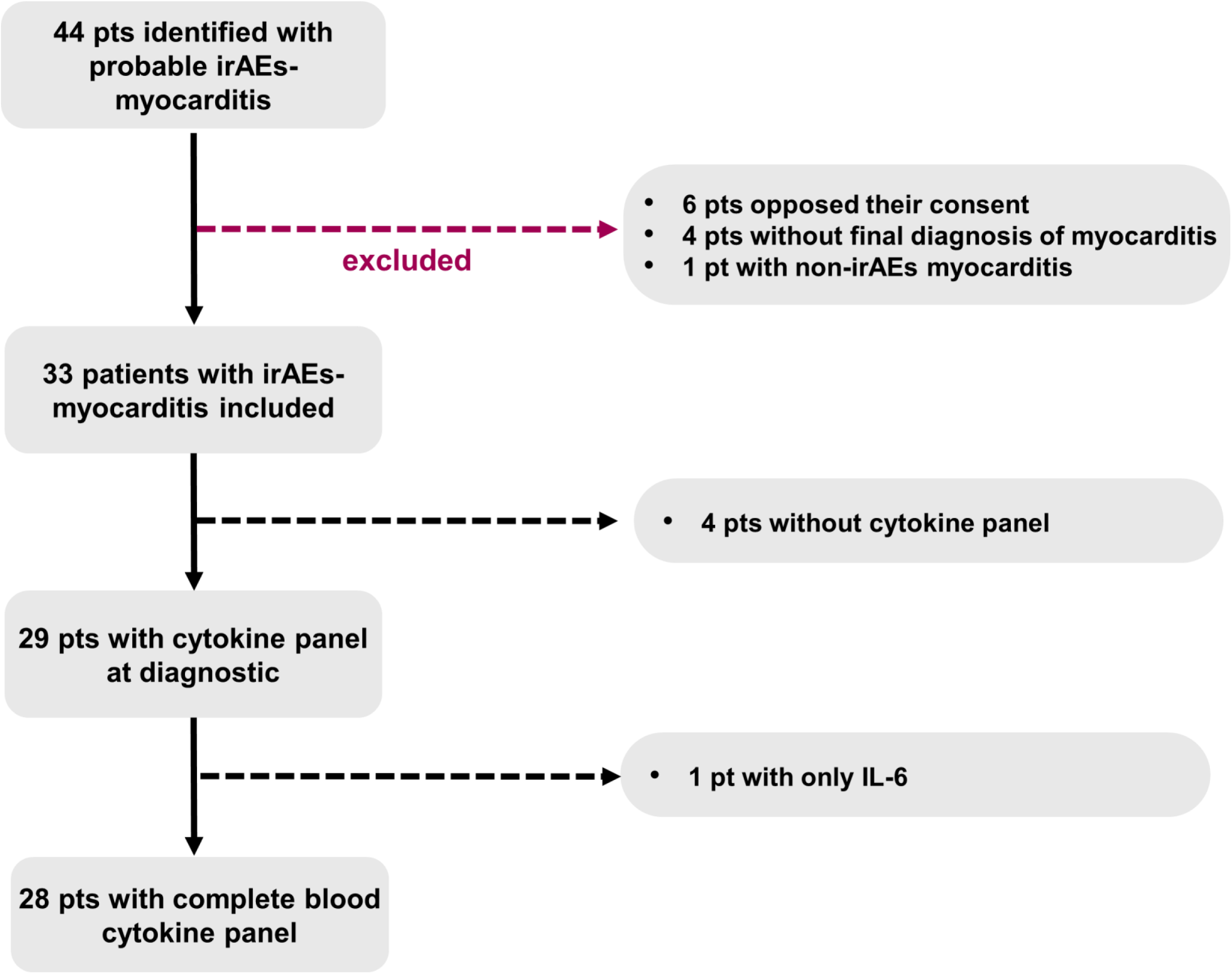
Gating strategy for mass cytometry. Complete gating strategy identify the main population with the list of antibodies and coupled antibodies.

**Supplementary Figure 3:**
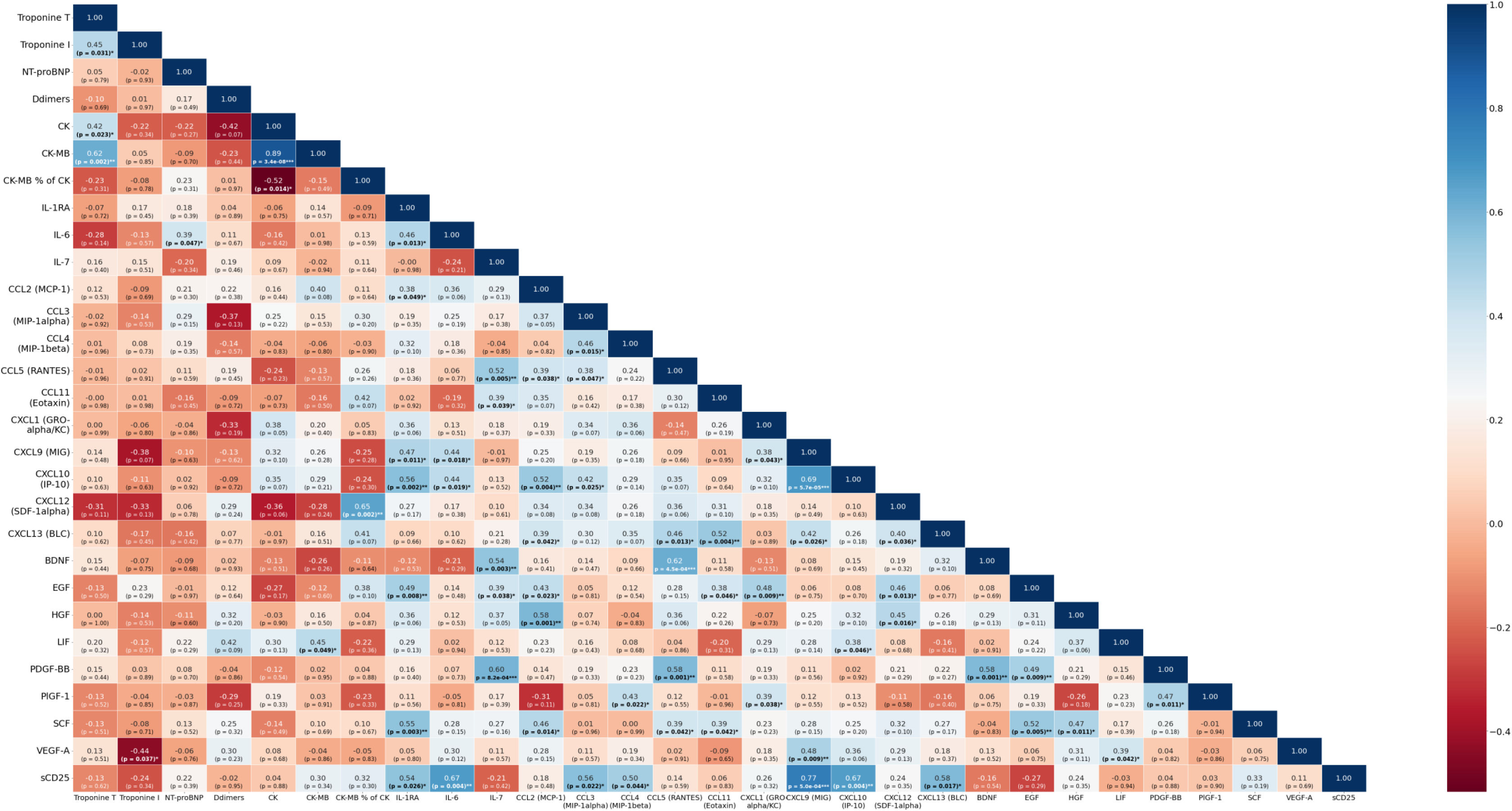
Correlation between cytokine and biological biomarkers in ICI-My. Spearman Correlation Matrix. Correlation map plotted using significance levels for the Spearman test performed with relevant serum biomarker data from all ICI-My patients studied across all grades. Positive correlations are shown in shaded blue and negative correlations are shown in shaded red. Correlations with a p-value ≥ 0.05 are not considered significant and are left blank. Color intensities are proportional to the correlation coefficients. On the right side of the correlogram, the color legend shows the correlation coefficients and corresponding colors.

**Supplementary Figure 4.**
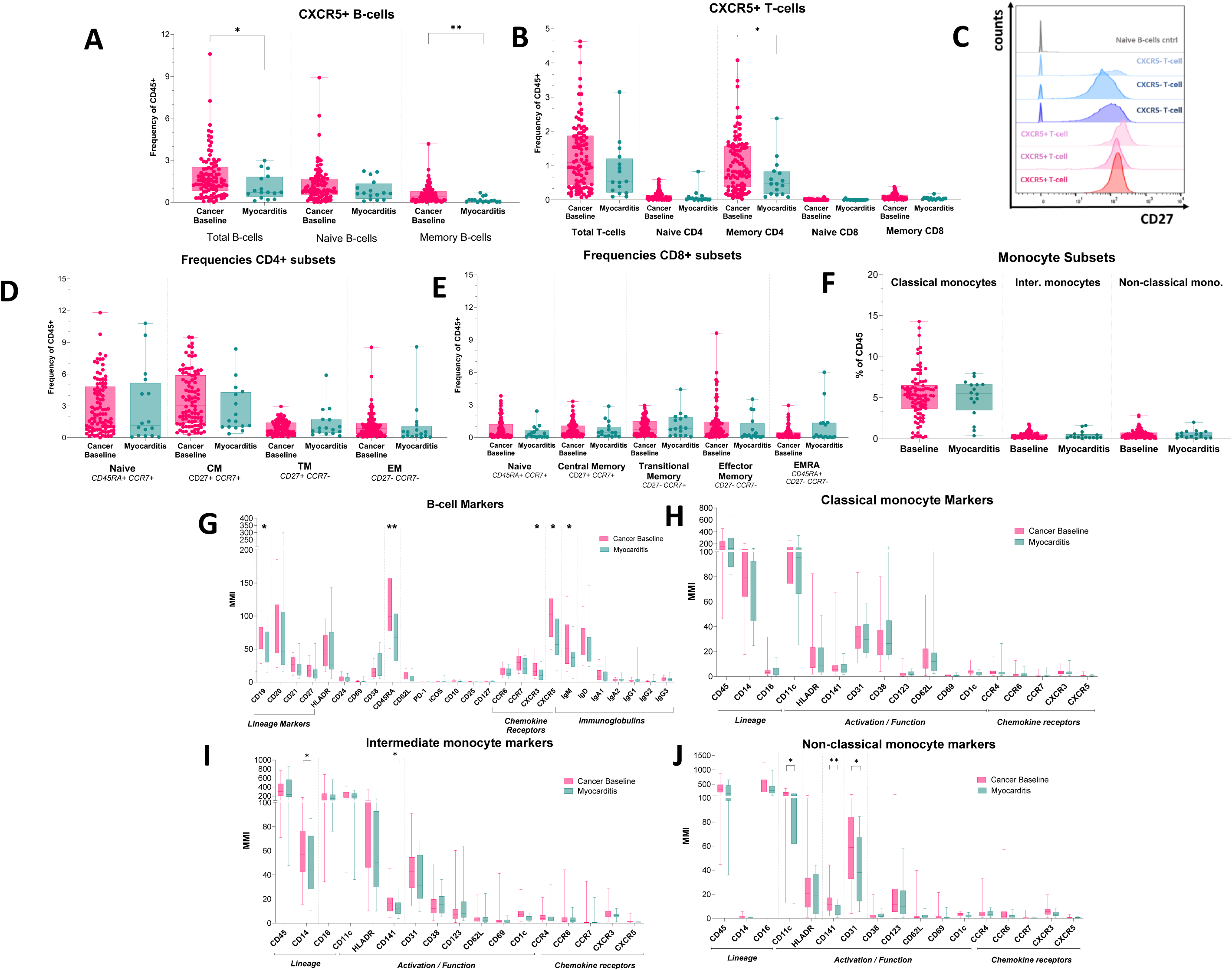
Distribution and phenotypes of circulating immune cell subsets in at myocarditis onset. A) Frequencies of CXCR5+ B-cells as total B-cells (CD19+CD20+), naive (IgD+CD27-) and memory (total – naive) in the circulation of cancer baseline and myocarditis patients, B) Frequencies of CXCR5+ T-cells as total T-cells (all CD3+), CD4/CD8 naive (CCR7+CD45RA+), CD4/CD8 memory (CD45RO+), C) Representative histograms showing CD27 expression in CXCR5+ (red) and CXCR5-(blue) T-cells in 3 representative cancer baseline patients. Negative control in grey (naive B-cells), D-E) Frequencies of CD4 and CD8 T-cell subsets as % of CD45+ cells, F) Frequencies of monocytes subsets (Classical, CD14+CD16-/Intermediate, CD14+CD16+ / Non-classical, CD14-CD16+) as % of CD45+, G) Marker expression on total B-cells (CD19+CD20+), expressed as mean metal intensity (MMI), H-J) Marker expression on classical, intermediate and non-classical monocytes expressed as MMI. Data were obtained from 16 patients at ICI-My diagnosis and compared with baseline levels from 97 cancer patients. Statistical significance was assessed using the Mann-Whitney U-Test, with significance levels as *p<0.05, **p<0.01.

**Supplementary Figure 5.**
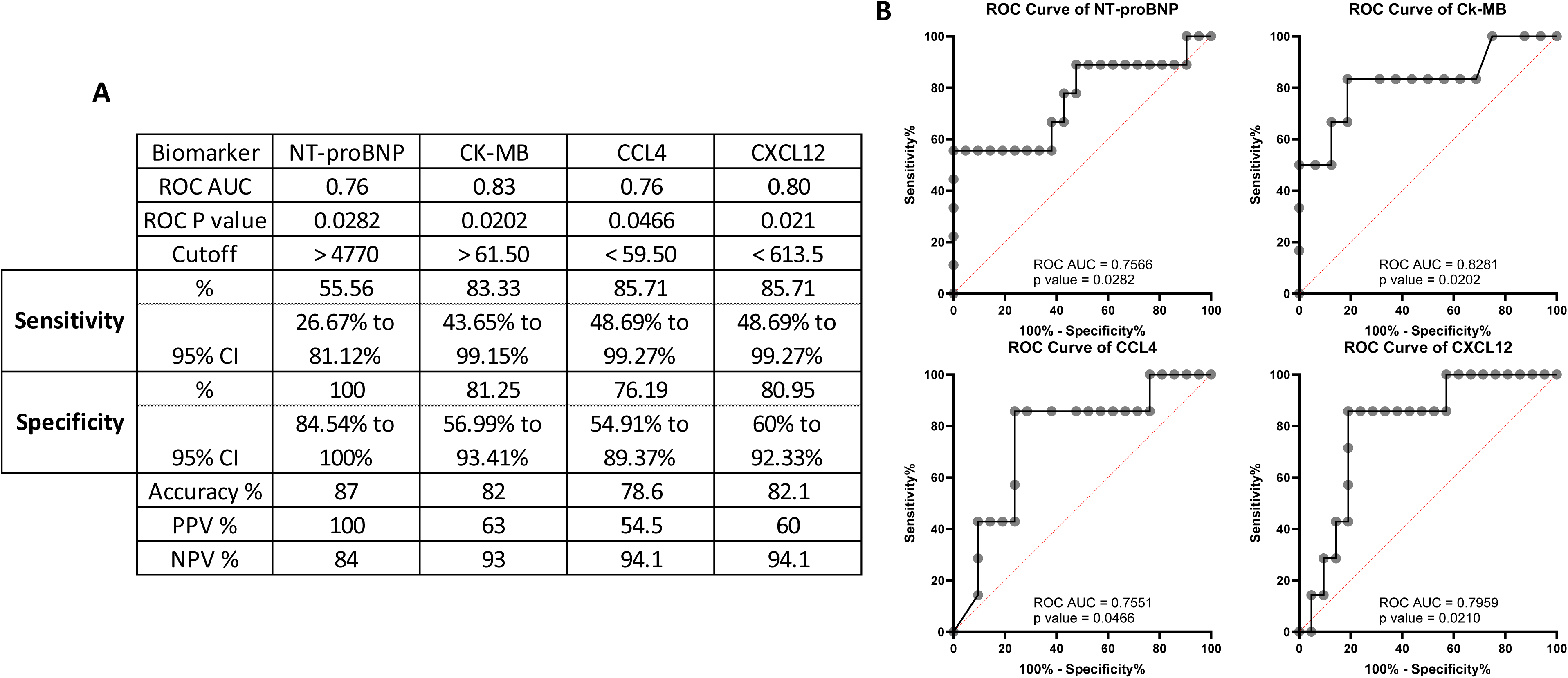
Comparison of biological profiles in low- and high-grade myocarditis. (A) Performance metrics include area under the curve (AUC), cutoff values, sensitivity, specificity, and predictive values for NT-proBNP, CK-MB, CCL4 and CXCL12 to distinguish low-grade (n=21) and high-grade (n=7) ICI-My. (B) Receiver operating characteristic (ROC) analysis of each significant marker’s discriminative ability to distinguish between low-grade (n=21) and high-grade (n=7) ICI-My. Confidence intervals for sensitivity and specificity were calculated using the Wilson-Brown method, optimal cutoff using the maximal Youden index.

**Supplementary Figure 6.**
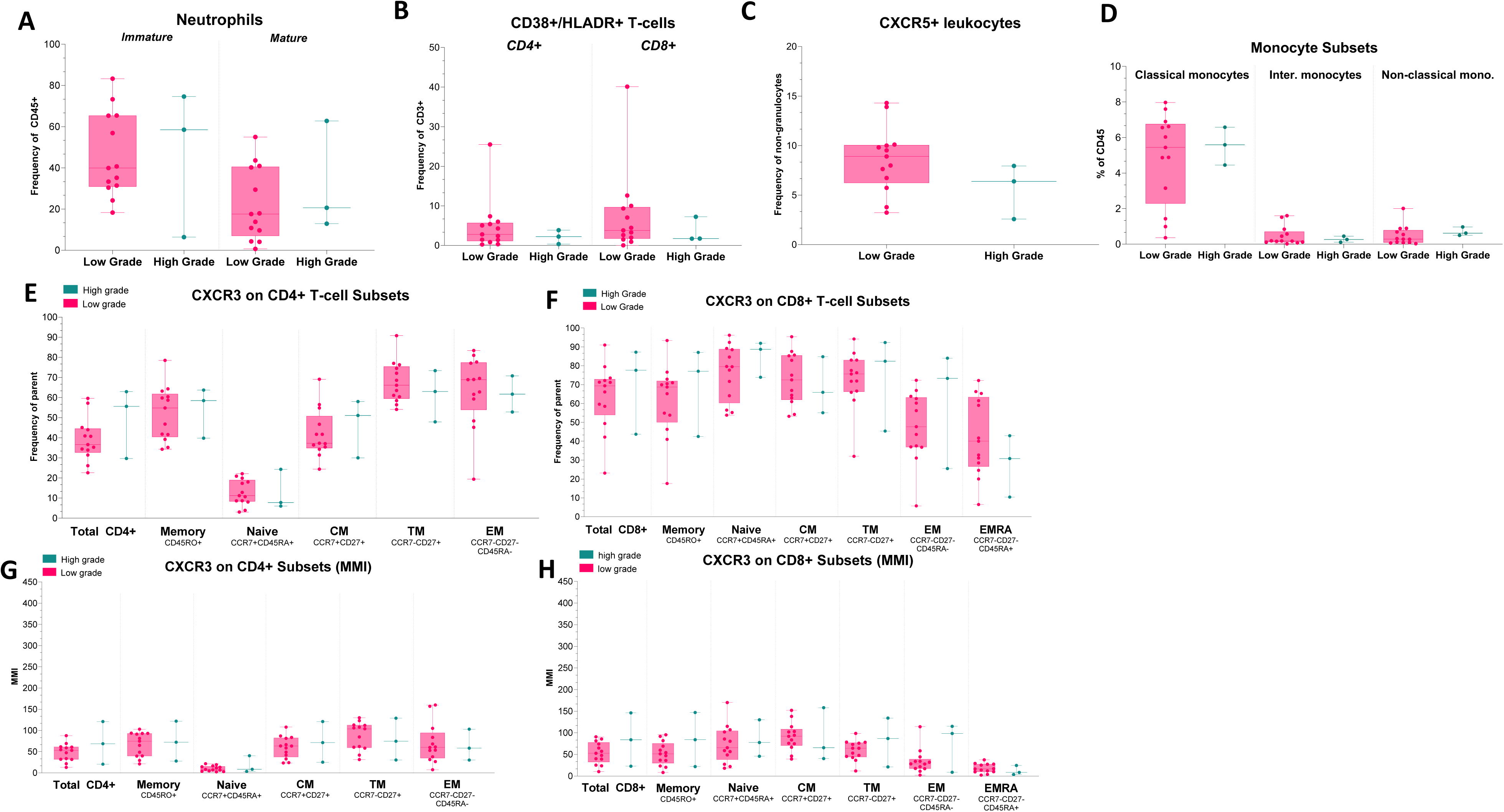
Comparison of distribution and phenotypes of circulating immune cell populations in low- and high-grade myocarditis. (A–G) Frequencies and phenotypic characteristics of circulating immune cell populations in myocarditis patients (low-grade: n=13; high-grade: n=3) compared to baseline levels from 98 cancer patients. A) Frequencies of circulating immature (CD10–) and mature (CD10+) neutrophils expressed as a percentage of CD45+ cells, B) Frequencies of activated T cells (CD38+ HLA-DR+) within CD4+ and CD8+ T-cell subsets, expressed as a percentage of total CD3+ cells, C) Frequency of CXCR5+ leukocytes expressed as a percentage of CD66b– cells, D) Frequencies of circulating immature monocytes subsets expressed as a percentage of CD45+ cells, E) Frequencies of CXCR3+ cells within CD8+ T-cell subsets, F) Frequencies of CXCR3+ cells within CD4+ T-cell subsets, G-H) CXCR3 expression measured by mean metal intensity (MMI) on CD8+ and CD4+ T-cell subsets. Data were obtained from 17 patients at ICI-My diagnosis and compared with baseline levels from 98 cancer patients. Statistical significance was determined using the Mann-Whitney U-test, with *P<0.05, **P<0.01, ***P<0.001, and ****P<0.0001.

**Supplementary Figure 7.**
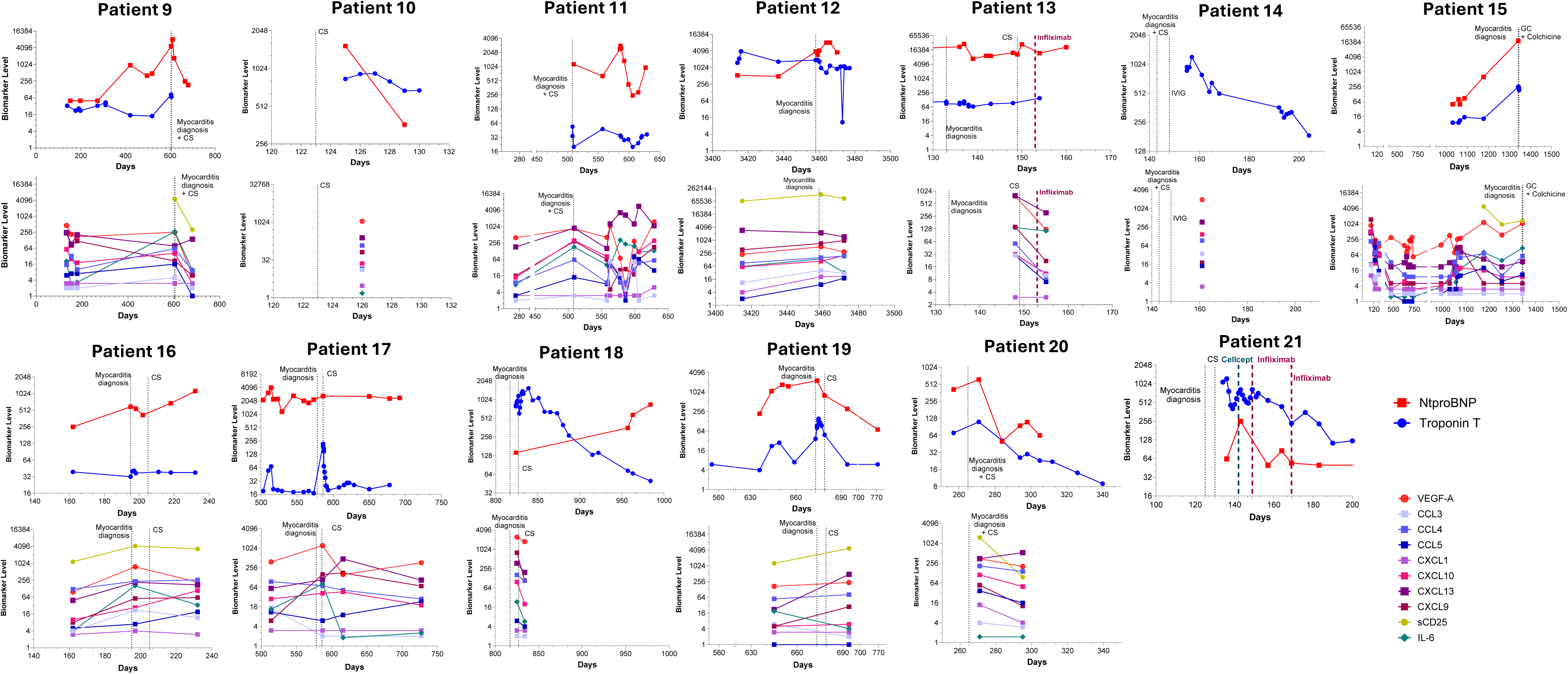
Longitudinal biomarker changes and evidence of biological resolution in ICI-Myocarditis patients treated with corticosteroids (CS) alone or in combination with other immunosuppressive (IS) agents, without tocilizumab (TCZ). Each panel represents an individual patient (n=16) receiving either CS only or a combination of CS with other IS therapies, excluding TCZ. Longitudinal trajectories of key cardiac biomarkers (cTnT [ng/L], NT-proBNP [ng/L]) and corresponding changes in selected cytokines, chemokines, and growth factors are depicted over time. The date of myocarditis onset and the timing of immunosuppressive treatments are indicated on each patient’s panel, illustrating how these interventions influence biomarker profiles and potentially contribute to biological resolution. Abbreviations: CS, corticosteroids.

**Supplementary Figure 8.**
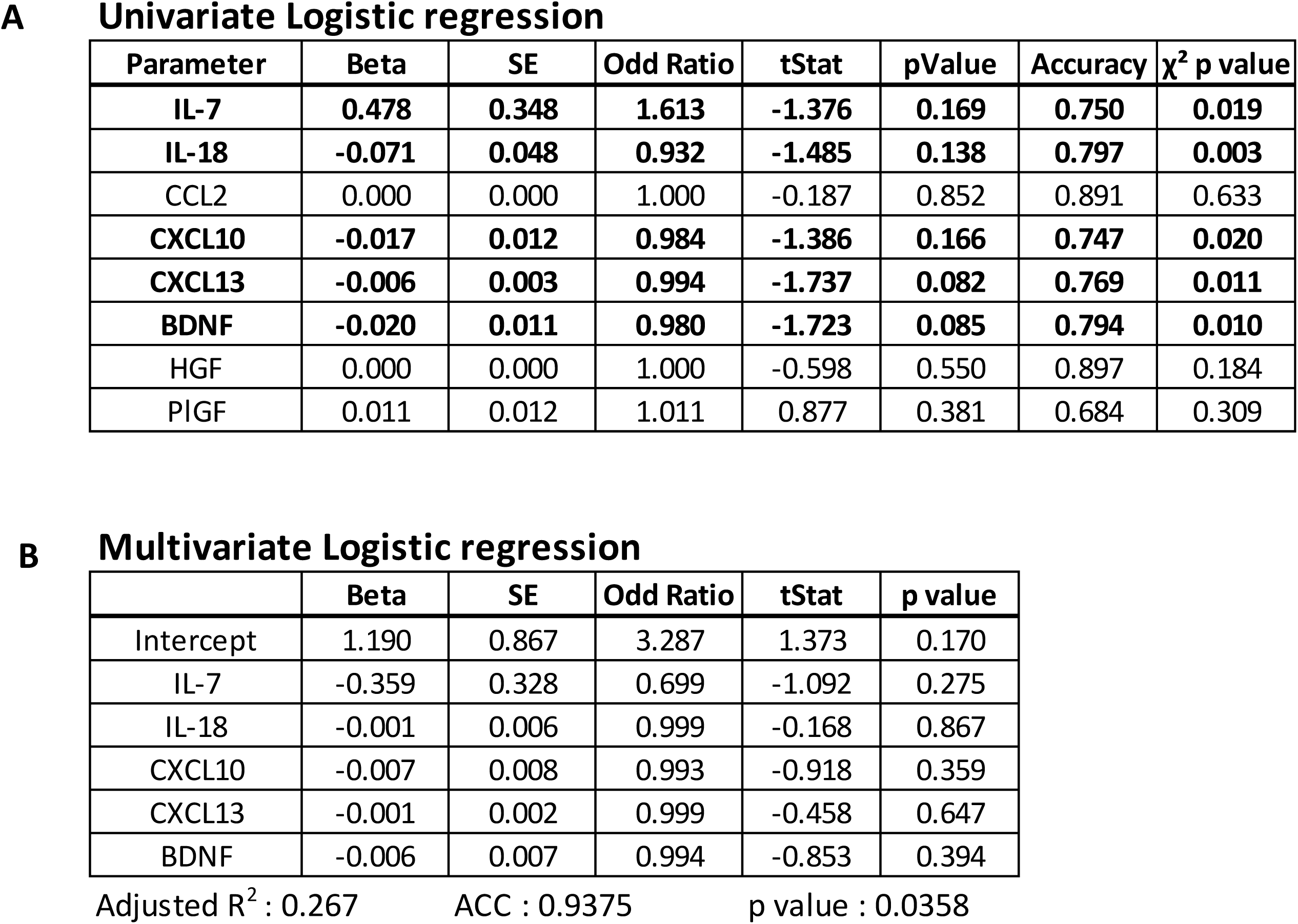
Univariate and Multivariate logistic regression predicting need for immunosuppressive treatment. Cytokines, Chemokines, and growth factor data for untreated (n=8) and immunosuppressive therapy treated patients (n=20) have been evaluated for their potential to be predictive of treatment requirement. Tables report beta coefficient, standard error, odd ratio, t stat, p value, accuracy of the classification, and chi square p value of the individual model or multivariate model for univariate (A) or multivariate models (B).

**Supplementary Figure 9.**
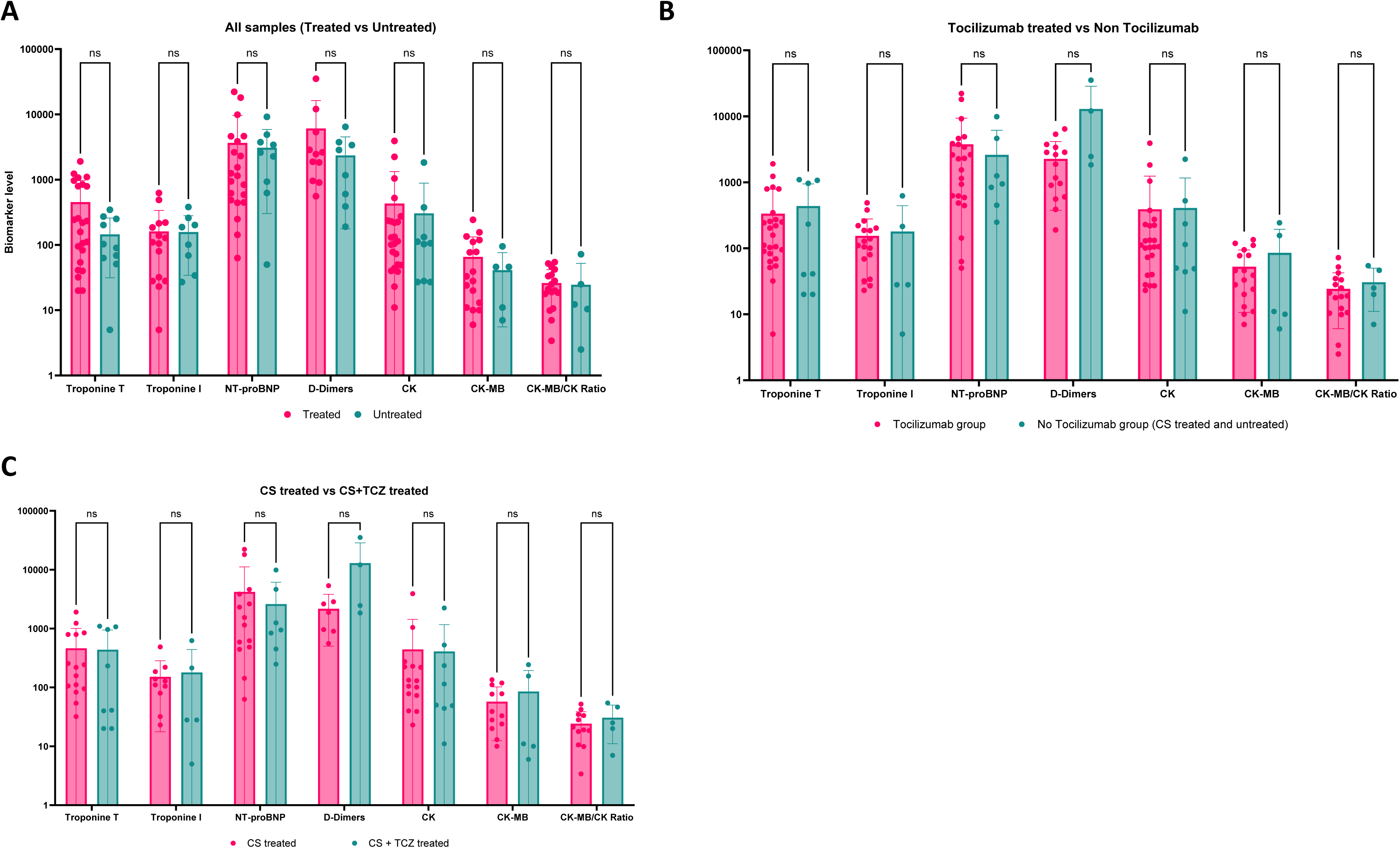
Comparison of classical biomarkers in patients regarding their need for therapy. Classical biomarkers in ICI-My patients have been compared in 3 groups, (A) receiving any immunosuppressive treatment (n=23) or no immunosuppressive treatment (n=10), (B) all patients non receiving tocilizumab (n=25) or receiving tocilizumab (n=8) (C) receiving corticosteroids (n=15) or receiving corticosteroid plus tocilizumab (n=8). Comparison was performed on serum cardiac troponin T (cTnT) (A n=33, B n=33, C n=23), serum cardiac troponin I (cTnI) (A n=23, B n=23, C n=15), NT-proBNP (A n=30, B n=30, C n=21), D-dimers (A n=19, B n=19, C n=11), Total creatine kinase (CK) (A n=32, B n=32, C n=23), Creatine kinase-MB (CK-MB) (A n=22, B n=22, C n=17) and CK-MB/CK ratio (A n=22, B n=22, C n=17) in high and low grade ICI-My diagnosis

Supplementary Table 1: Table summarizing N, Mean, Minimum, 25% Percentile, Median, 75% Percentile, 90% Percentile and Maximum for all cytokines, chemokines and growth factor depending of the ICI-My grade.

Supplementary Table 2: Cohort description including clinical data, treatment, and classical biomarkers levels.

Supplementary Table 3: Clinical characteristics of irCRS patients receiving Tocilizumab treatment (n=7).

