## Supplementary Table 3 for "Immune profiling identifies predictive biomarkers and highlights the potential efficacy of IL-6R blockade in checkpoint inhibitor–related myocarditis"

| | N=8 | % within cohort |
|---|---|---|
| **Tumor type** | | |
| Melanoma | 3 | 37% |
| Lung | 2 | 24% |
| Breast | 1 | 13% |
| Kidney | 1 | 13% |
| Prostate | 1 | 13% |
| **ICI treatment** | | |
| Anti-CTLA4/PD1 | 6 | 75% |
| Anti-PD1 | 2 | 25% |
| **Associated irAEs** | | |
| Colitis | 4 | 50% |
| Myocarditis-myositis overlap syndrome | 1 | 13% |
| CRS | 1 | 13% |
| Cholangio-hepatitis | 1 | 13% |
| **Myocarditis grading at diagnosis** | | |
| Low Grade (G1-G2) | 5 | 63% |
| High Grade (G3-G4) | 3 | 38% |
| **Previous immunosupression therapy** | | |
| High-dose steroids (HDS) | 8 | 100% |
| Infliximiab (IFX) | 2 | 25% |
| Mycophenolic mofetil (MMF) | 1 | 13% |
| **Tocilizumab therapy** | | |
| Mortality related to myocarditis | 0 | 0% |
| Clinical improvement | 8 | 100% |
| Biological and cytokine improvement (n=7) | 7 | 100% |
| CS tapering | 8 | 100% |
| 1 Dose of tocilizumab administered | 4 | 50% |
| 2 Dose of tocilizumab administered | 1 | 13% |
| 3 Dose of tocilizumab administered | 3 | 38% |
